# Semaglutide is associated with improved breast cancer survival, lower metastatic burden, and a dose–survival relationship uncoupled from weight-loss magnitude

**DOI:** 10.64898/2026.04.23.26351609

**Authors:** Karthik Murugadoss, AJ Venkatakrishnan, Venky Soundararajan

**Author notes:** Correspondence: Venky Soundararajan.

## Abstract

Metabolic dysfunction is increasingly recognized as a risk factor for poor outcomes in breast cancer, but whether incretin-based therapies confer survival benefit beyond weight loss remains unresolved. Using a federated electronic health record platform spanning nearly 29 million patients, we evaluated breast cancer survival after semaglutide and tirzepatide initiation in routine care. In 1:1 propensity-matched pooled-comparator analyses, semaglutide was associated with improved overall survival versus metformin, sodium–glucose cotransporter 2 (SGLT2) inhibitor, and dipeptidyl peptidase 4 (DPP4) inhibitor users, with 54 deaths among 2,433 semaglutide users (2.2%) versus 395 deaths among 2,433 comparators (16.2%) over 24 months (log-rank P < 0.001). Tirzepatide showed a favorable survival association relative to pooled anti-diabetic comparators that did not meet statistical significance (P = 0.24), with 3 deaths among 220 users (1.4%) versus 64 deaths among 220 comparators (29.1%). In a head-to-head propensity-score–matched comparison, overall survival did not differ significantly between semaglutide- and tirzepatide-treated patients with pre-existing breast cancer (2,117 per arm; P = 0.12). In semaglutide-treated patients alive and observable at the 1-year landmark, higher maximum dose achieved was significantly associated with lower post-landmark mortality (P = 0.034), with an event rate of approximately 1.0% in the high-dose group (≥1.7 mg) versus approximately 4.5% in the low-dose group (0.25–1.0 mg). Despite a linear dose–weight loss relationship for semaglutide, however, weight-loss strata did not separate survival outcomes (global P = 0.22). In tirzepatide-treated patients alive and observable at the same landmark, neither maximum dose achieved nor weight-loss strata separated post-landmark survival (P = 0.98 and P = 0.50, respectively). Structured EHR and AI-based clinical-note analyses further showed significantly lower frequency of documented metastatic disease in semaglutide-treated patients relative to pooled anti-diabetic comparators, including any metastasis (7.0% versus 15.0%, rate ratio 0.5, P < 0.001), bone metastasis (1.0% versus 5.2%, rate ratio 0.2, P < 0.001), and liver, lung, or brain metastases (all P < 0.001). LLM-derived cause-of-death extraction further showed a 60% lower relative proportion of cancer-associated deaths in semaglutide-treated patients (19% of ascertainable deaths) than in matched pooled anti-diabetic comparators (47% of ascertainable deaths), with comparator deaths more often attributed to cancer progression involving metastatic breast cancer, leptomeningeal carcinomatosis, and cancer-driven organ failure. Overall, this study demonstrates that semaglutide use in patients with pre-existing breast cancer is associated with a dose-correlated but weight-loss independent improvement in overall survival. These findings motivate prospective trials of GLP-1 receptor agonists in breast cancer across various stages and treatment settings.

## Introduction

Obesity, metabolic dysfunction and breast cancer intersect through insulin resistance, inflammation, altered adipokine signalling and endocrine disruption. In breast-cancer survivors, higher body mass index has been associated with worse overall and breast-cancer-specific survival, and post-diagnosis weight trajectories have likewise been linked to subsequent mortality, highlighting the importance of host metabolic state after diagnosis^1,2^.

Against this backdrop, incretin-based therapies have transformed the treatment landscape for obesity and type 2 diabetes. Once-weekly semaglutide produced substantial and sustained weight loss in STEP 1, semaglutide reduced major adverse cardiovascular events in SELECT, and tirzepatide improved clinical outcomes in obesity-associated heart failure with preserved ejection fraction in SUMMIT^3–5^. At the same time, tirzepatide yielded greater weight reduction than semaglutide in a head-to-head obesity trial, reinforcing the need to distinguish effects driven primarily by body-weight change from those linked more broadly to pharmacological exposure^6^.

Whether GLP-1 pathway therapies influence cancer outcomes remains unresolved. In a large observational study of patients with type 2 diabetes, GLP-1 receptor agonists were associated with lower incidence of several obesity-associated cancers compared with insulin, but were not associated with reduced postmenopausal breast-cancer risk and did not show a clear advantage over metformin for overall cancer prevention^7^. In adults with obesity, target-trial emulation likewise suggested lower overall cancer incidence among GLP-1 receptor agonist users, although the signal was driven mainly by selected tumour types rather than breast cancer specifically^8^. By contrast, among older adults with cancer and type 2 diabetes, GLP-1 receptor agonist use was associated with lower all-cause mortality than DPP4 inhibitor use, including within breast-cancer subgroups, suggesting that survival and cancer incidence may not align^9^.

Breast-cancer–specific clinical data on incretin therapy remain limited. A recent clinical series evaluating GLP-1 receptor agonist use in patients with breast cancer described clinically meaningful weight loss over follow-up, but did not directly resolve whether outcome associations in this setting are better explained by weight reduction itself or by drug-exposure–related biology^3,10^. This distinction is important because GLP-1 receptor agonists exert pleiotropic effects beyond weight loss, including changes in glycaemia, cardiometabolic physiology and systemic inflammatory state^3–5,10^.

Here we used the nference federated electronic health record platform, spanning nearly 29 million patients, to evaluate survival outcomes among patients with prior breast cancer who subsequently initiated semaglutide or tirzepatide in routine care. We first compared post-initiation survival for semaglutide and tirzepatide versus active glucose-lowering comparators, then integrated structured EHR data with AI-derived clinical-note curation and LLM-based cause-of-death extraction to determine whether observed survival associations aligned more closely with drug exposure, metastatic burden, or achieved weight loss. Among semaglutide-treated patients alive and observable at a 1-year landmark, we next tested whether post-landmark survival tracked more closely with maximum dose achieved, treatment persistence, or weight-loss magnitude. Finally, we incorporated tumor-genomic survival analyses of GLP1R and GIPR alterations to place the clinical findings in the context of receptor-pathway biology.

## Results

### Semaglutide is associated with improved survival compared to other anti-diabetic medications in patients with previously diagnosed breast cancer

To assess whether GLP-1 receptor agonist therapy was associated with altered survival in patients with previously diagnosed breast cancer, we assembled 1:1 propensity matched cohorts of patients with diagnosed breast cancer who subsequently initiated semaglutide or another anti-diabetic medication (metformin, sodium-glucose cotransporter 2 [SGLT2] inhibitors, or dipeptidyl peptidase 4 [DPP4] inhibitors). In the primary semaglutide analysis, 2,433 patients were matched per arm. The baseline demographic and clinical characteristics used for propensity matching were well balanced between the cohorts, including age, baseline body mass index (BMI) and weight, type 2 diabetes mellitus status, time since breast cancer diagnosis, time since most recent oncology therapy, and prior oncologic therapy exposures (**Table 1**). Median follow-up was approximately 421 days [IQR 182 to 769] in semaglutide users and 665 days [IQR 223 to 1,531] in pooled comparators. Over 24 months of follow-up, the semaglutide cohort was associated with lower mortality than the pooled comparator cohort with 54 deaths among 2,433 semaglutide users (2.2%) versus 395 deaths among 2,433 comparators (16.2%) (p < 0.001; **Figure 1a**; **Table 1**). AI-based curation of clinical notes in the 2-year pre-index window identified higher rates of documented breast cancer across multiple stages in the comparator cohort, including Stages I (144/2433 [5.9%] vs. 91/2433 [3.7%], p<0.001), II (86/2433 [3.5%] vs. 50/2433 [2.1%], p=0.002), III (60/2433 [2.5%] vs. 28/2433 [1.2%], p<0.001), and IV (41/2433 [1.7%] vs. 24/2433 [1.0%], p=0.034); semaglutide-treated patients had higher rates of invasive ductal breast carcinoma Stage IA documentation (45/2433 [1.9%] vs. 17/2433 [0.7%], p<0.001). These differences reflect rates of clinical documentation in the pre-index period and may not necessarily represent the true underlying disease burden in either cohort.

**Figure 1.**
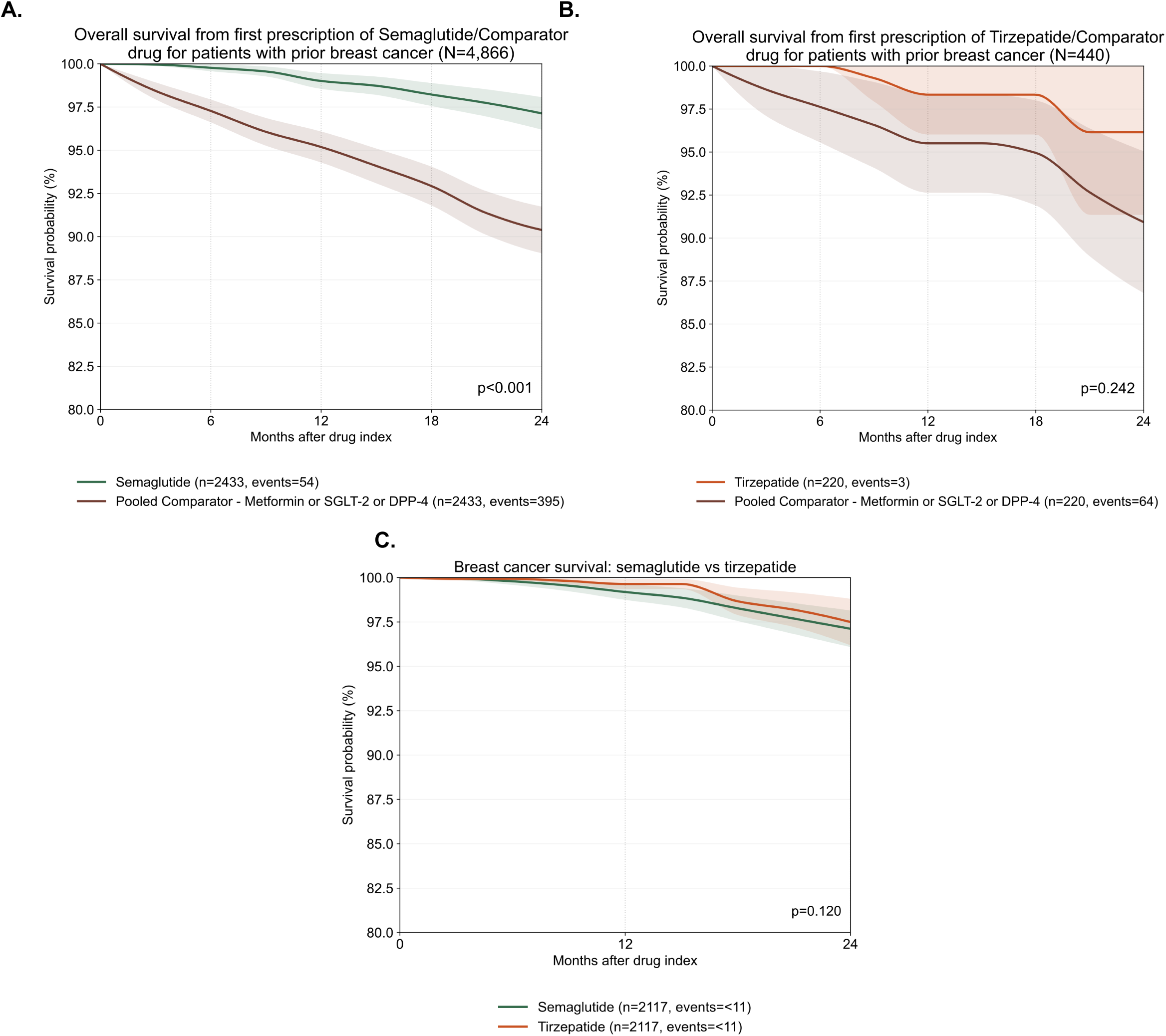
Overall survival from first prescription of semaglutide or tirzepatide versus pooled comparator glucose-lowering comparators in patients with prior breast cancer. **(A)** Kaplan-Meier estimates of overall survival from first prescription of semaglutide versus a pooled comparator cohort comprising metformin, sodium-glucose cotransporter 2 (SGLT2) inhibitors, and dipeptidyl peptidase 4 (DPP4) inhibitors. Semaglutide users (n = 2,433) showed higher survival probability over the 24-month display window than pooled comparators (n = 2,433), with 24-month Kaplan-Meier event probabilities of 2.9% versus 9.6% (log-rank P < 0.001). **(B)** Kaplan-Meier estimates of overall survival from first prescription of tirzepatide versus the same pooled-comparator framework. Tirzepatide users (n = 220) showed a numerically favorable but non-significant pattern relative to pooled comparators (n = 220), with 24-month Kaplan-Meier event probabilities of 3.8% versus 9.1% (P = 0.242). **(C)** Kaplan-Meier estimates of overall survival from first prescription of semaglutide versus tirzepatide in a 1:1 propensity-score–matched cohort of patients with prior breast cancer. Semaglutide users (n = 2,117) and tirzepatide users (n = 2,117) showed no significant difference in survival over the 24-month display window, respectively (P = 0.120). Shaded areas indicate 95% confidence intervals.

**Table 1.** Demographics and baseline characteristics of semaglutide-treated versus the pooled comparator cohort, including all variables that were considered for 1:1 propensity matching.

| **VARIABLE (1:1 propensity matched)** | **Semaglutide** | **Pooled comparator** |
| --- | --- | --- |
| N patients | 2433 | 2433 |
| Age at index, mean (SD) | 62.2 (11.1) | 62.4 (11.4) |
| Baseline BMI, mean (SD) | 33.9 (6.0) | 34.0 (6.7) |
| Baseline weight, mean (SD) | 92.0 (18.7) | 92.3 (21.1) |
| Index year, mean (SD) | 2023 (1.3) | 2020 (4.8) |
| Days from qualifying prior breast cancer to index, mean (SD) | 2193.7 (1842.7) | 2089.9 (1850.1) |
| Days from most recent oncology treatment to index, mean (SD) | 2653.5 (6536.5) | 2523.3 (6067.7) |
| Female sex, n (%) | 2419 (99.4%) | 2401 (98.7%) |
| T2DM at baseline, n (%) | 449 (18.5%) | 460 (18.9%) |
| Any prior oncology treatment, n (%) | 2418 (99.4%) | 2412 (99.1%) |
| Prior endocrine therapy, n (%) | 1673 (68.8%) | 1667 (68.5%) |
| Prior chemotherapy, n (%) | 666 (27.4%) | 608 (25.0%) |
| Prior HER2-directed therapy, n (%) | 225 (9.2%) | 218 (9.0%) |
| Prior CDK4/6 inhibitor, n (%) | 101 (4.2%) | 81 (3.3%) |
| Prior PARP inhibitor, n (%) | 13 (0.5%) | 3 (0.1%) |
| Prior checkpoint immunotherapy, n (%) | 41 (1.7%) | 33 (1.4%) |
| Prior other targeted therapy, n (%) | 24 (1.0%) | 29 (1.2%) |
| Follow up days, median [IQR] | 421 [182, 769] | 665 [223, 1,531] |

### Tirzepatide is associated with a numerically lower but statistically non-significant reduction in mortality in patients with previously diagnosed breast cancer

The same protocol was followed to derive 1:1 propensity-matched cohorts of breast cancer patients who subsequently initiated tirzepatide versus another anti-diabetic medication (N = 220 each). Again, the baseline characteristics that were used for propensity matching were well balanced between these groups (**Table 2**). While the risk of death over 24 months of follow-up was numerically lower in the tirzepatide-treated cohort, with 24-month Kaplan-Meier event probabilities of 3.8% versus 9.1% and corresponding incidence rates of 1.48 versus 5.36 deaths per 100 person-year, this difference was not statistically significant (p = 0.24) (**Figure 1b**).

**Table 2.** Demographics and baseline characteristics of the tirzepatide-treated versus the pooled comparator cohort, including all variables that were considered for 1:1 propensity matching.

| **VARIABLE** | **Tirzepatide** | **Pooled comparator** |
| --- | --- | --- |
| N patients | 220 | 220 |
| Age at index, mean (SD) | 63.9 (15.0) | 64.3 (9.7) |
| Baseline BMI, mean (SD) | 35.3 (7.2) | 31.4 (6.2) |
| Baseline weight, mean (SD) | 98.1 (25.0) | 93.8 (18.7) |
| Index year, mean (SD) | 2024 (0.9) | 2023 (4.9) |
| Days from qualifying prior breast cancer to index, mean (SD) | 2739.1 (2871.0) | 1378.4 (1641.9) |
| Days from most recent oncology treatment to index, mean (SD) | 2120.0 (3201.0) | 2200.0 (3100.0) |
| Female sex, n (%) | 219 (99.5%) | 218 (99.1%) |
| T2DM at baseline, n (%) | 23 (10.5%) | 25 (11.4%) |
| Any prior oncology treatment, n (%) | 213 (96.8%) | 213 (96.8%) |
| Prior endocrine therapy, n (%) | 147 (66.8%) | 147 (66.8%) |
| Prior chemotherapy, n (%) | 57 (25.9%) | 57 (25.9%) |
| Prior HER2-directed therapy, n (%) | 19 (8.6%) | 19 (8.6%) |
| Prior CDK4/6 inhibitor, n (%) | <11 (N/A) | <11 (N/A) |
| Prior PARP inhibitor, n (%) | <11 (N/A) | <11 (N/A) |
| Prior checkpoint immunotherapy, n (%) | <11 (N/A) | <11 (N/A) |
| Prior other targeted therapy, n (%) | <11 (N/A) | <11 (N/A) |

### Head-to-head propensity-score–matched analysis shows no significant overall-survival difference between semaglutide and tirzepatide

In a head-to-head propensity-score–matched analysis of patients with prior breast cancer, 2,713 semaglutide users and 2,368 tirzepatide users were identified, of whom 2,535 and 2,117, respectively, were eligible for matching; 2,117 patients per arm were then matched 1:1 (**Table 3**). Matching was performed on age at index, baseline BMI, baseline weight, index year, time from qualifying prior breast cancer diagnosis to index, sex, and baseline type 2 diabetes status. Time from most recent oncology treatment to index and prior oncology treatment class were not included in the matching model because their inclusion resulted in insufficient matched counts. In the matched cohort, overall survival (2.9% vs 2.5% respectively) was not significantly different between semaglutide and tirzepatide (P = 0.120) (**Figure 1c**).

**Table 3.**
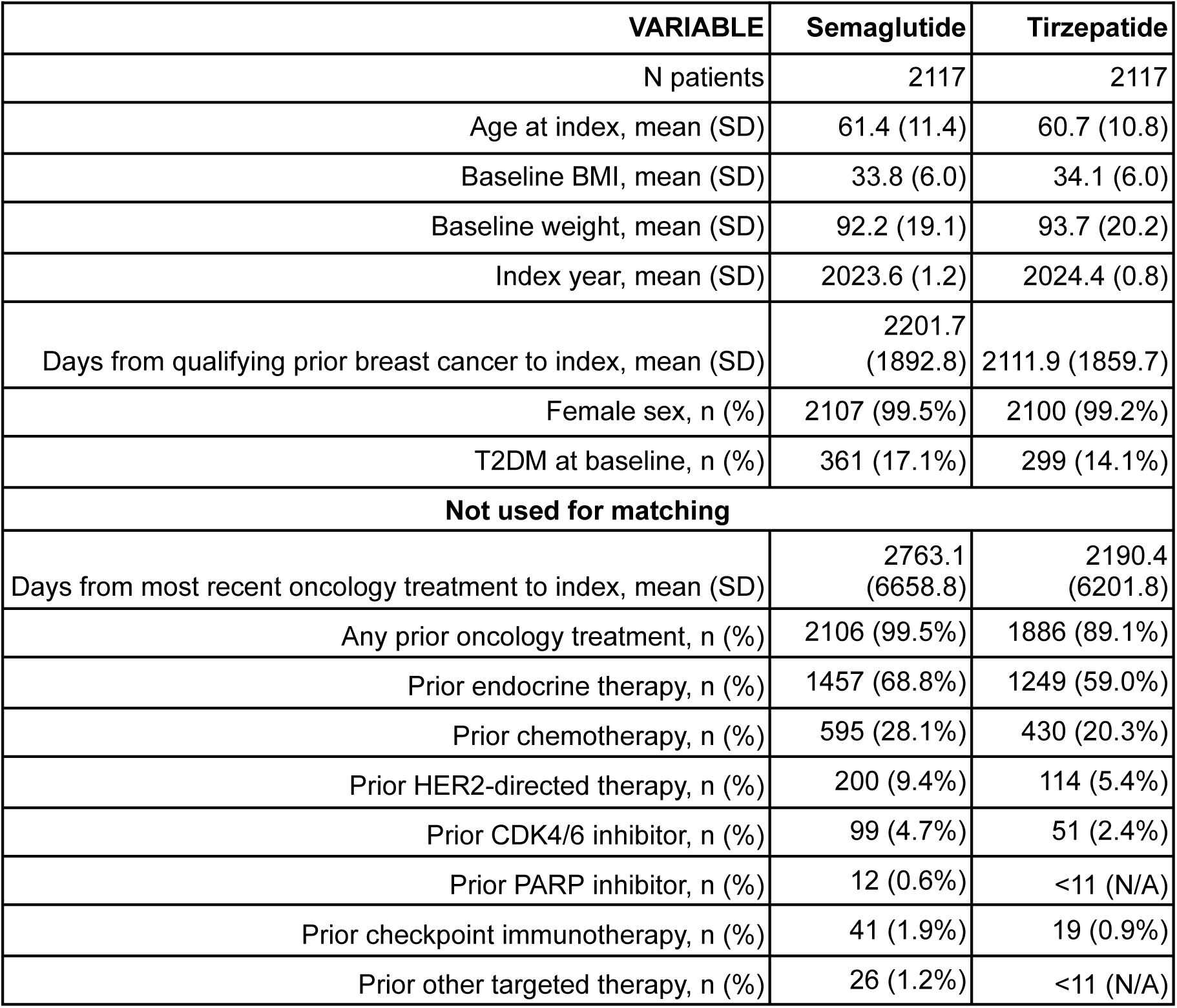
Demographics and baseline characteristics of the semaglutide-treated versus tirzepatide-treated versus the pooled comparator cohort, including all variables that were considered for 1:1 propensity matching. Prior treatment was not included for matching.

| **VARIABLE** | **Semaglutide** | **Tirzepatide** |
| --- | --- | --- |
| N patients | 2117 | 2117 |
| Age at index, mean (SD) | 61.4 (11.4) | 60.7 (10.8) |
| Baseline BMI, mean (SD) | 33.8 (6.0) | 34.1 (6.0) |
| Baseline weight, mean (SD) | 92.2 (19.1) | 93.7 (20.2) |
| Index year, mean (SD) | 2023.6 (1.2) | 2024.4 (0.8) |
| Days from qualifying prior breast cancer to index, mean (SD) | 2201.7<br>(1892.8) | 2111.9 (1859.7) |
| Female sex, n (%) | 2107 (99.5%) | 2100 (99.2%) |
| T2DM at baseline, n (%) | 361 (17.1%) | 299 (14.1%) |
| **Not used for matching** |  |  |
| Days from most recent oncology treatment to index, mean (SD) | 2763.1<br>(6658.8) | 2190.4<br>(6201.8) |
| Any prior oncology treatment, n (%) | 2106 (99.5%) | 1886 (89.1%) |
| Prior endocrine therapy, n (%) | 1457 (68.8%) | 1249 (59.0%) |
| Prior chemotherapy, n (%) | 595 (28.1%) | 430 (20.3%) |
| Prior HER2-directed therapy, n (%) | 200 (9.4%) | 114 (5.4%) |
| Prior CDK4/6 inhibitor, n (%) | 99 (4.7%) | 51 (2.4%) |
| Prior PARP inhibitor, n (%) | 12 (0.6%) | <11 (N/A) |
| Prior checkpoint immunotherapy, n (%) | 41 (1.9%) | 19 (0.9%) |
| Prior other targeted therapy, n (%) | 26 (1.2%) | <11 (N/A) |

### Concordant structured EHR and AI-derived clinical note analyses show significantly lower metastatic burden with semaglutide than matched pooled anti-diabetic drugs

In propensity-matched cohorts of patients with prior breast cancer who subsequently initiated semaglutide (n = 2,433) or a pooled comparator therapy comprising metformin, SGLT-2 inhibitors, and DPP-4 inhibitors (n = 2,433), metastatic burden post-semaglutide or comparator initiation was consistently lower in the semaglutide cohort than in comparators. In comparators, structured ICD-coded diagnoses identified significantly higher rates of lung metastasis (<11 vs. 51 patients, <0.5% vs. 2.1%, p<0.001), pleural metastasis (<11 vs. 17 patients, <0.5% vs. 0.7%, p=0.010), skin metastasis (<11 vs. 15 patients, <0.5% vs. 0.6%, p=0.025), and metastatic cancer (<11 vs. 15 patients, <0.5% vs. 0.6%, p=0.025; **Table 4**). AI-based unstructured clinical notes curation corroborated and extended these findings, identifying any metastasis (including but not limited to breast cancer metastasis itself) in 171 (7%) vs. 365 (15%) patients (rate ratio 0.5, p<0.001), with particularly striking differences for bone metastasis (24 [1%] vs. 127 [5.2%], rate ratio 0.2, p<0.001), liver metastasis (<11 vs. 62 [2.5%], p<0.001), lung metastasis (<11 vs. 59 [2.4%], p<0.001) and brain metastasis (<11 vs. 47 [1.9%], p<0.001; **Table 4**). Before treatment initiation, AI-based extraction identified any metastasis in 177/2,433 (7.3%) semaglutide-treated versus 281/2,433 (11.5%) comparator patients, and bone metastasis in 15 (0.6%) semaglutide-treated versus 71 (2.9%) comparator patients, while ICD-coded lung metastasis was present in <11 semaglutide-treated versus 23 (0.9%) comparator patients (**Table 4**). These baseline differences indicate that part of the post-treatment metastatic separation may reflect residual pre-existing imbalance, although the persistence of concordant differences across structured and AI-derived analyses supports further evaluation in designs that more tightly balance baseline metastatic burden.

**Table 4.** Post-treatment metastatic disease burden in semaglutide-treated versus comparator cohorts. Prevalence of metastatic disease diagnoses in the post-index period across propensity-matched semaglutide (n=2,433) and pooled comparator (n=2,433) cohorts of patients with previously diagnosed breast cancer. Metastatic diagnoses were identified using two complementary approaches: structured ICD-10 diagnosis codes queried from the electronic health record, and AI-based natural language processing of unstructured clinical notes using GPT-OSS (20 billion parameter model). Rate ratios are expressed as semaglutide relative to comparator; values reported as upper bounds (<) reflect semaglutide patient counts below the institutional privacy threshold of 11. These findings are exploratory and hypothesis-generating.

| Metastasis Finding | Semaglutide pre-tx, n (%) | Comparator, pre-tx, n (%) | Semaglutide, post-tx, n (%) | Comparator post-tx, n (%) | Rate Ratio (post-tx) | P-value (post-tx) |
| --- | --- | --- | --- | --- | --- | --- |
| **Structured ICD Codes** |  |  |  |  |  |  |
| Lung metastasis [C78.00, C78.01, C78.02] | <11 (<0.5%) | 23 (0.9%) | <11 (<0.5%) | 51 (2.1%) | <0.20 | <0.001 |
| Pleural metastasis [C78.2] | - | - | <11 (<0.5%) | 17 (0.7%) | <0.59 | 0.01 |
| Skin metastasis [C79.2] | <11 (<0.5%) | <11 (<0.5%) | <11 (<0.5%) | 15 (0.6%) | <0.67 | 0.025 |
| Metastatic cancer (unspecified site) [C79.9] | <11 (<0.5%) | 13 (0.5%) | <11 (<0.5%) | 15 (0.6%) | <0.67 | 0.025 |
| **LLM Extraction** |  |  |  |  |  |  |
| Any metastasis | 177 (3.1%) | 281 (11.5%) | 171 (7.0%) | 365 (15.0%) | 0.47 | <0.001 |
| Bone metastasis | 15 (0.6%) | 71 (2.9%) | 24 (1.0%) | 127 (5.2%) | 0.19 | <0.001 |
| Liver metastasis | <11 (<0.5%) | 13 (0.5%) | <11 (<0.5%) | 62 (2.5%) | <0.16 | <0.001 |
| Lung metastasis | - | - | <11 (<0.5%) | 59 (2.4%) | <0.17 | <0.001 |
| Brain metastasis | - | - | <11 (<0.5%) | 47 (1.9%) | <0.21 | <0.001 |

### LLM-derived cause-of-death analyses suggest fewer cancer-associated deaths in semaglutide-treated patients than in matched pooled anti-diabetic comparators

Overall mortality was substantially lower in the semaglutide cohort than in comparators (54/2,433, 2.2%, versus 395/2,433, 16.2%). Among deaths with an ascertainable cause, cancer progression, including metastatic breast cancer, leptomeningeal carcinomatosis, and cancer-driven organ failure, was the dominant category in the comparator cohort, accounting for approximately 47% of deaths (186 cases), whereas cancer progression including metastatic and leptomeningeal disease accounted for a substantially smaller share of deaths in the semaglutide cohort (∼19%; rate ratio 0.39, P < 0.001; **Table 5**). By contrast, cardiac events and sepsis/septic shock represented larger proportions of the much smaller semaglutide death pool (∼30% and ∼22%, respectively). This divergence in cause-of-death profiles is directionally consistent with the lower metastatic burden observed in the semaglutide cohort. Given the substantially smaller number of tirzepatide-treated patients and deaths, an analogous cause-of-death comparison for tirzepatide could not be meaningfully performed at this time.

**Table 5.** Cause of death in semaglutide-treated versus comparator cohorts. Causes of death among all patients who died in the propensity-matched semaglutide (n=54 deaths, 2.2%) and comparator (n=395 deaths, 16.2%) cohorts. Causes of death were extracted from unstructured clinical notes using a large language model and harmonized into six high-level categories. Patient counts for individual categories in the semaglutide cohort are simulated from LLM-derived proportions applied to the total death count and should be interpreted as approximate. These findings are exploratory and hypothesis-generating.

| Cause of Death | Semaglutide, n (%) | Comparator, n (%) | Rate Ratio | p-value |
| --- | --- | --- | --- | --- |
| Cancer Progression / Metastasis | <11 (19%) | 186 (47%) | 0.39 | <0.001 |
| Cardiac Events | 16 (30%) | 55 (14%) | 2.13 | 0.003 |
| Sepsis / Septic Shock | 12 (22%) | 43 (11%) | 2.04 | 0.017 |
| Respiratory Failure | <11 (11%) | 63 (16%) | 0.70 | 0.351 |
| Hemorrhage / Stroke | <11 (7%) | 20 (5%) | 1.46 | 0.474 |
| Other | <11 (11%) | 28 (7%) | 1.57 | 0.296 |

### In patients with pre-existing breast cancer, higher attained semaglutide dose is associated with improved all-cause mortality after one year of semaglutide exposure

To assess whether post-landmark survival aligned more closely with maximum dose achieved or with weight-loss magnitude, we stratified the semaglutide-treated population based on one measure of exposure (the maximum dose achieved) during the first year after semaglutide initiation (“pre-landmark interval”). As expected, higher maximum dose of semaglutide was associated with increased magnitude of weight loss during this interval, with approximately 3.31% greater maximum weight loss per 1 mg increase in semaglutide dosage (**Figure 2a**). Among the patients who were still at-risk at the 1-year landmark, the post-landmark survival was then assessed between strata defined by either maximum dose achieved or maximum weight loss during the pre-landmark interval. Post-landmark all-cause mortality was lower in patients who had reached a higher maximum dose of semaglutide (≥1.7 mg; N = 444) compared to those with a lower maximum dose (0.25-1.0 mg; N = 955) (p = 0.034; **Figures 3a-b**). On the other hand, post-landmark all-cause mortality was similar between the weight-loss stratified cohorts (p = 0.22; **Figures 3c-d**). Thus, although a higher achieved semaglutide dose during the first year was associated with greater weight loss, post-landmark survival among 1-year survivors aligned more closely with maximum dose than with weight loss magnitude.

**Figure 2.**
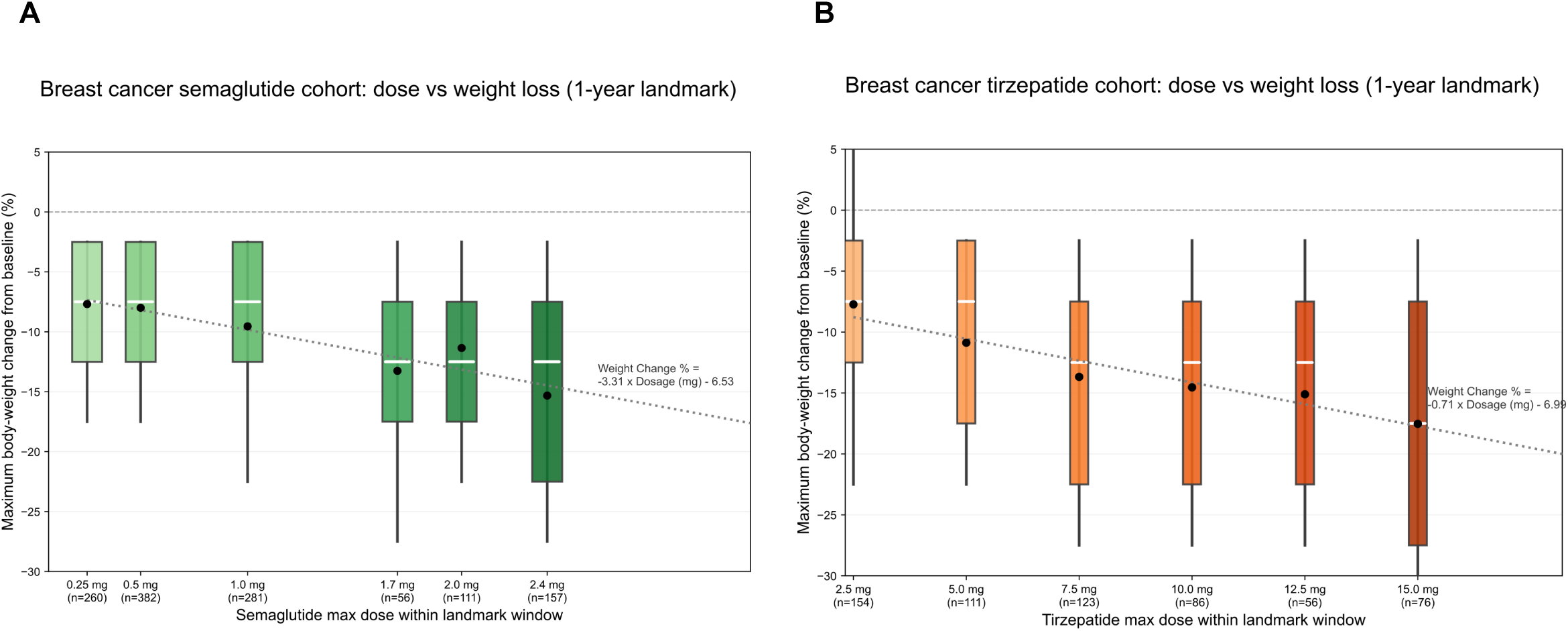
Maximum dose achieved is associated with greater body-weight reduction within the 1-year landmark window. **(A)** Distribution of maximum body-weight change from baseline across semaglutide maximum dose groups in patients with prior breast cancer who were alive and observable at the 1-year landmark. Dose groups were 0.25 mg (n = 260), 0.5 mg (n = 382), 1.0 mg (n = 281), 1.7 mg (n = 56), 2.0 mg (n = 111) and 2.4 mg (n = 157). The fitted relationship between maximum dose achieved and body-weight change was: weight change (%) = −3.31 × dosage (mg) − 6.53. **(B)** Analogous analysis for tirzepatide maximum dose groups: 2.5 mg (n = 154), 5.0 mg (n = 111), 7.5 mg (n = 123), 10.0 mg (n = 86), 12.5 mg (n = 56) and 15.0 mg (n = 76). The fitted relationship was: weight change (%) = −0.71 × dosage (mg) − 6.99. In both panels, boxes indicate the interquartile range, center lines indicate medians, whiskers indicate the full observed range, black dots indicate group means, and dotted lines indicate linear fits across dose groups. Negative values denote reductions in body weight relative to baseline.

**Figure 3.**
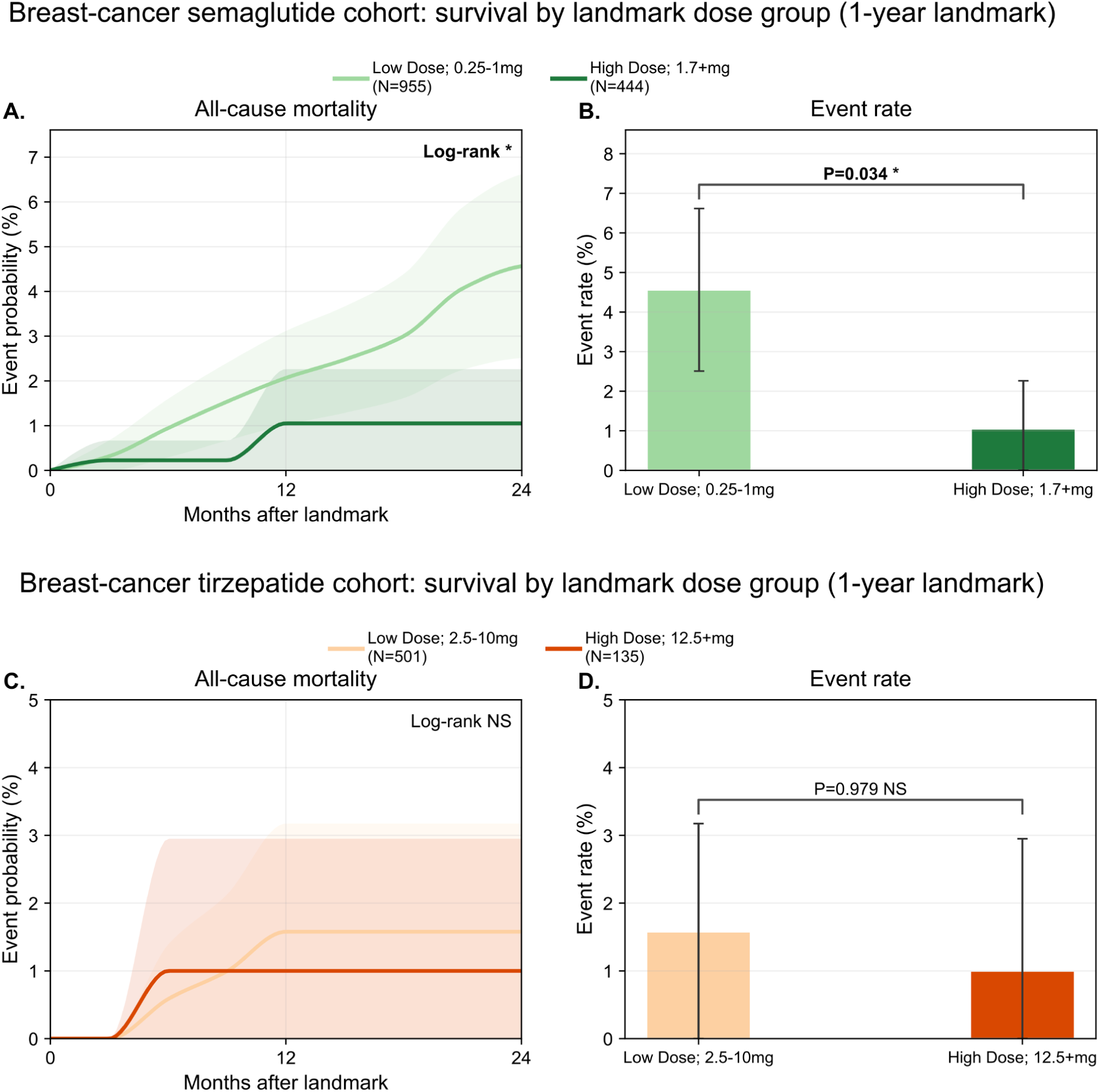
Post-landmark survival by maximum dose achieved within the 1-year landmark window. **(A-B)** Semaglutide landmark-dose analysis in patients with prior breast cancer who were alive and observable at 1 year after treatment initiation. a, Kaplan–Meier estimates of post-landmark all-cause mortality comparing lower-dose semaglutide (0.25–1.0 mg; n = 955) with higher-dose semaglutide (≥1.7 mg; n = 444). b, Corresponding event-rate comparison. Higher maximum dose achieved was associated with lower post-landmark mortality, with event rates of approximately 4.5% versus 1.0% in the low- and high-dose groups, respectively (log-rank P = 0.034). **(C-D)** Analogous tirzepatide landmark-dose analysis. c, Kaplan–Meier estimates of post-landmark all-cause mortality comparing lower-dose tirzepatide (2.5–10 mg; n = 501) with higher-dose tirzepatide (≥12.5 mg; n = 135). d, Corresponding event-rate comparison. Post-landmark mortality did not differ significantly by maximum dose achieved, with event rates of approximately 1.5% versus 1.0% in the low- and high-dose groups, respectively (P = 0.979). In Kaplan–Meier panels, shaded areas indicate 95% confidence intervals. In event-rate panels, bars show group event rates and error bars indicate 95% confidence intervals.

### Tirzepatide landmark analyses do not show a significant dose- or weight-loss–stratified survival difference

In an analogous landmark-based analysis of tirzepatide, higher maximum dose of tirzepatide achieved during the pre-landmark period was similarly associated with body weight reduction, with approximately 0.71% greater maximum weight loss per 1 mg dose increase (**Figure 2b**). Among the patients who were still at-risk at the 1-year landmark, there was no significant difference in post-landmark survival between patients who had achieved a lower maximum dose (2.5-10 mg; N = 501) compared to those who had achieved a higher maximum dose (12.5-15 mg; N = 135) (p = 0.98, **Figures 4a-b**). Similarly, post-landmark survival did not differ between the subcohorts of tirzepatide-treated patients stratified by maximum weight loss during the pre-landmark interval (p = 0.50; **Figures 4c-d**).

**Figure 4.**
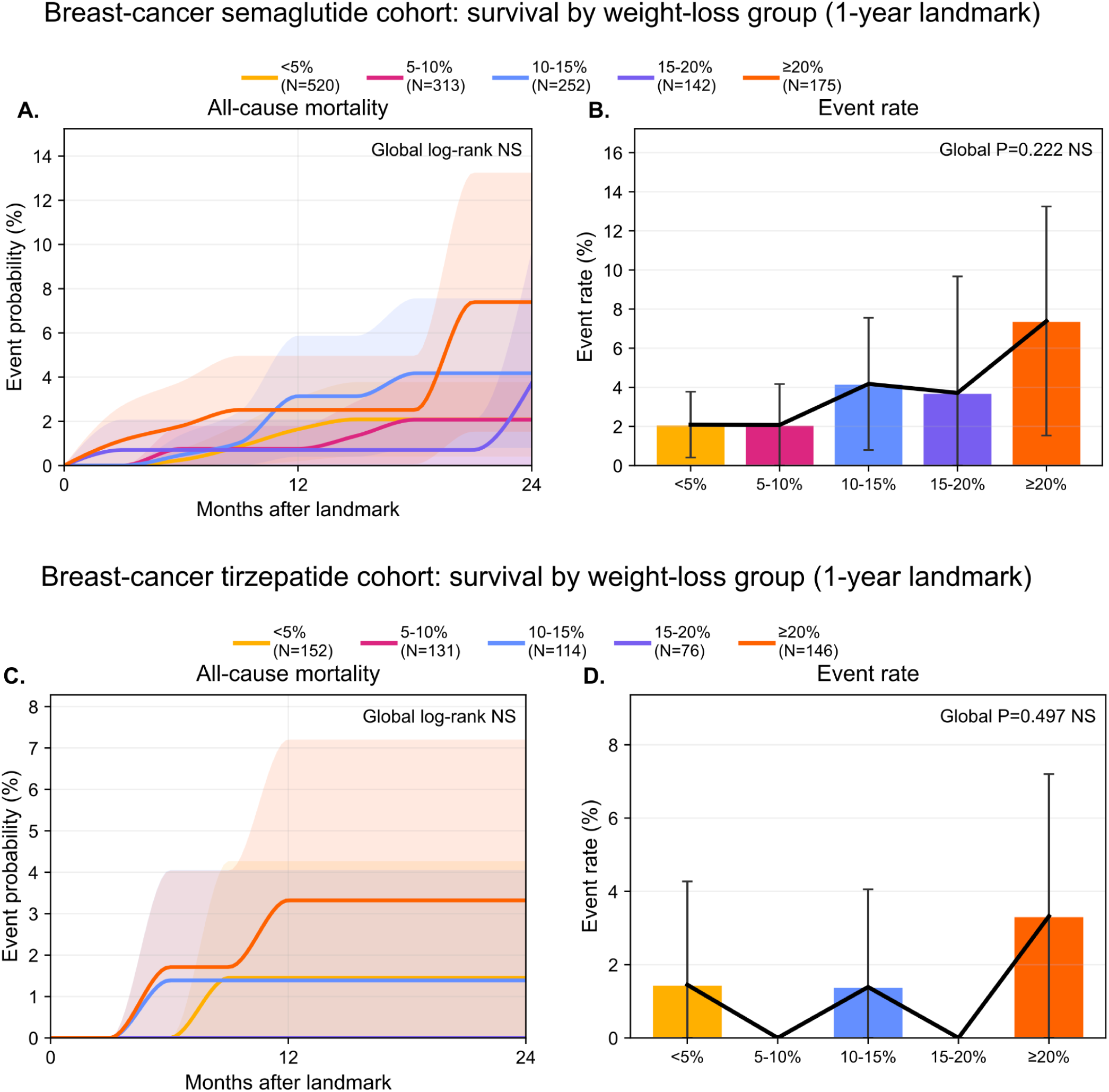
Post-landmark survival by weight-loss category within the 1-year landmark window. **(A-B)** Semaglutide landmark weight-loss analysis in patients with prior breast cancer who were alive and observable at 1 year after treatment initiation. a, Kaplan–Meier estimates of post-landmark all-cause mortality across weight-loss strata defined within the pre-landmark window: <5% (n = 520), 5–10% (n = 313), 10–15% (n = 252), 15–20% (n = 142) and ≥20% (n = 175). b, Corresponding event-rate comparison. Post-landmark survival did not differ significantly across semaglutide weight-loss groups (global log-rank P = 0.222). **(C-D)** Analogous tirzepatide landmark weight-loss analysis. c, Kaplan–Meier estimates of post-landmark all-cause mortality across tirzepatide weight-loss strata: <5% (n = 152), 5–10% (n = 131), 10–15% (n = 114), 15–20% (n = 76) and ≥20% (n = 146). d, Corresponding event-rate comparison. Post-landmark survival did not differ significantly across tirzepatide weight-loss groups (global P = 0.497). In Kaplan–Meier panels, shaded areas indicate 95% confidence intervals. In event-rate panels, bars show group event rates and error bars indicate 95% confidence intervals.

### Semaglutide-associated survival remained favorable when follow-up was anchored to oncology treatment initiation

To assess the potential impact of semaglutide as a priming agent for actively treated breast cancer, an additional analysis was performed in which breast cancer patients who received semaglutide prior to an oncology therapy were propensity-matched to those who received a different anti-diabetic medication prior to an oncology therapy. The risk of all-cause mortality, assessed from the start of the oncology therapy, was lower in the semaglutide cohorts than in each comparator cohort (**Figure 5**, **Table 6**). In the matched comparison of semaglutide versus metformin (n = 1,909 patients each), the incidence rates of death were 1.64 and 4.58 per 100 person-years, respectively (P < 0.001). These values were 3.15 versus 12.76 per 100 person-years for DPP4 inhibitors (n = 237 patients each; P = 0.002) and 2.12 versus 8.76 per 100 person-years for SGLT2 inhibitors (n = 1,089 patients each; P < 0.001).

**Figure 5.**
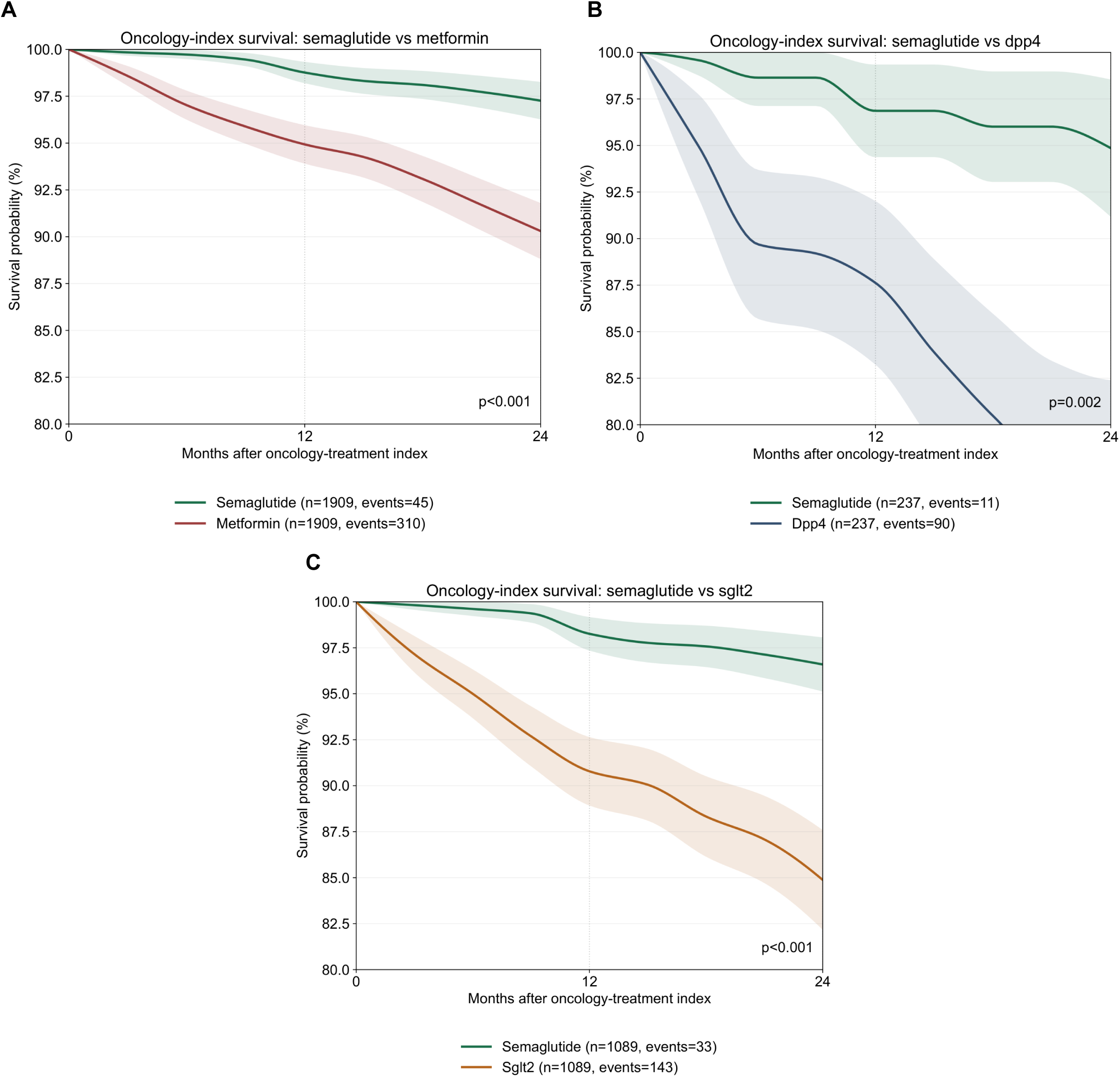
Semaglutide is associated with improved breast cancer survival across matched comparator cohorts when considering oncology treatment initiation as the index date. Kaplan–Meier estimates of overall survival in patients with breast cancer comparing semaglutide with matched comparator cohorts, with follow-up anchored at oncology-treatment index. **(A)** semaglutide versus metformin; **(B)** semaglutide versus DPP4 inhibitors; **(C)** semaglutide versus SGLT2 inhibitors. In the metformin-matched analysis, 1,909 patients were included per group and the 24-month Kaplan-Meier event probability was 2.7% in semaglutide users versus 9.7% in metformin users (P < 0.001). In the DPP4-matched analysis, 237 patients were included per group and the corresponding 24-month event probabilities were 5.1% versus 23.6% (P = 0.002). In the SGLT2-matched analysis, 1,089 patients were included per group and the corresponding 24-month event probabilities were 3.4% versus 15.1% (P < 0.001). Shaded areas indicate 95% confidence intervals.

**Table 6.** Demographics and baseline characteristics of the semaglutide-treated versus pooled comparator cohorts in the oncology-treatment-indexed analysis, including all variables that were considered for 1:1 propensity matching.

| VARIABLE | Semaglutide | Pooled comparator |
| --- | --- | --- |
| N patients | 1909 | 1909 |
| Age at index, mean (SD) | 62.3 (11.0) | 62.5 (11.4) |
| Baseline BMI, mean (SD) | 34.1 (6.1) | 34.2 (6.6) |
| Baseline weight, mean (SD) | 92.7 (18.8) | 92.6 (21.0) |
| Index year, mean (SD) | 2023.7 (1.2) | 2020.4 (4.5) |
| Days from qualifying prior breast cancer to index, mean (SD) | 2299.2 (1792.1) | 2213.1 (1801.5) |
| Days from most recent oncology treatment to index, mean (SD) | 56.0 (108.0) | 59.5 (120.7) |
| Female sex, n (%) | 1907 (99.9%) | 1895 (99.3%) |
| T2DM at baseline, n (%) | 367 (19.2%) | 380 (19.9%) |
| Any prior oncology treatment, n (%) | 1909 (100.0%) | 1909 (100.0%) |
| Prior endocrine therapy, n (%) | 1333 (69.8%) | 1329 (69.6%) |
| Prior chemotherapy, n (%) | 548 (28.7%) | 527 (27.6%) |
| Prior HER2-directed therapy, n (%) | 191 (10.0%) | 165 (8.6%) |
| Prior CDK4/6 inhibitor, n (%) | 84 (4.4%) | 80 (4.2%) |
| Prior PARP inhibitor, n (%) | 8 (0.4%) | 6 (0.3%) |
| Prior checkpoint immunotherapy, n (%) | 32 (1.7%) | 27 (1.4%) |
| Prior other targeted therapy, n (%) | 20 (1.0%) | 25 (1.3%) |
| Follow up days, median [IQR] (approx.) | 437 [210, 776] | 793 [341, 1,917] |

### Genomic and functional perturbation analyses of GLP1R and GIPR in breast cancer

To contextualize these clinical findings, we next assessed *GLP1R* and *GIPR* across public databases of tumor genomics and functional perturbation analyses. Across 30 individual studies including 4,726 total patients with genomic profiling (copy number alterations and/or mutations) accessed from the cBioPortal for Cancer Genomics^11–13^, *GLP1R* alterations were observed in 78 (1.7%) patients (**Table S2**). The majority of these alterations were a copy number alteration (CNA), including amplification in 66 of 3,724 (1.8%) profiled patients and deep deletions in 2 (0.05%) patients. There were *GLP1R* mutations observed in 12 of 2,311 (0.5%) profiled patients, without any pattern of recurrent hotspot mutations (**Figure S1A**). *GLP1R* alteration status was not associated with overall survival (p = 0.40, q = 0.80; **Figure 6a**), nor with progression-free survival (p = 0.63; q = 0.80) or disease-specific survival (p = 0.52; q = 0.80).

**Figure 6.**
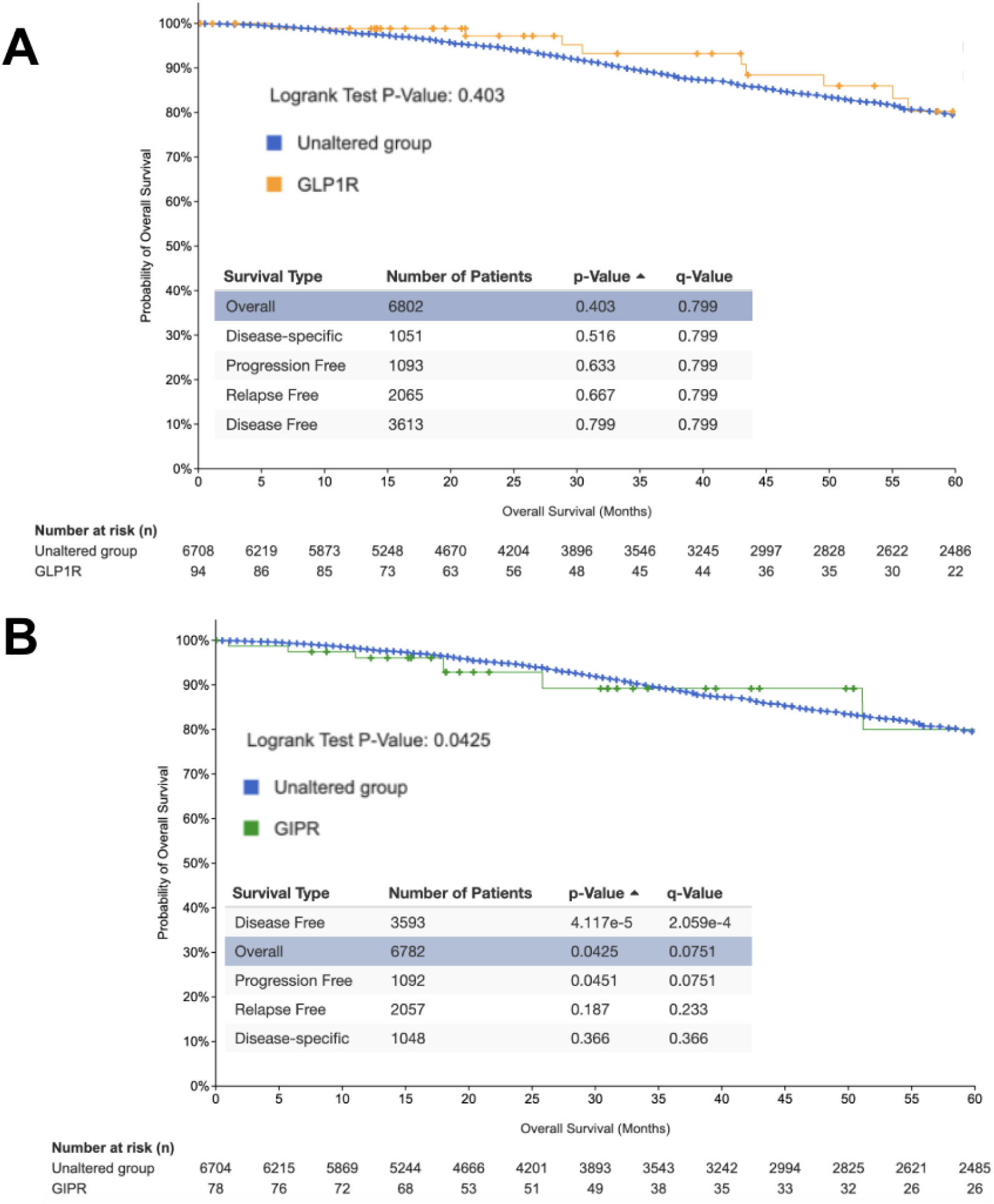
Overall survival in patients with *GLP1R*- or *GIPR*-altered breast cancer. **(A)** Kaplan–Meier estimates of overall survival comparing **(A)** *GLP1R*-altered or **(B)** *GIPR*-altered breast cancer samples with unaltered samples across the aggregated breast-cancer cohort. The survival curves shown depict overall survival. The statistics shown in the inset correspond to other time-to-event analyses including disease-specific, disease-free, progression-free, and relapse-free survival. Tick marks indicate censored observations. Data is sourced from https://www.cbioportal.org/results/comparison/survival?comparison_selectedGroups=%5B%2[...]plots_vert_selection=%7B%7D&plots_coloring_selection=%7B%7D (*GLP1R*) and https://www.cbioportal.org/results/comparison/survival?comparison_selectedGroups=%5B%2[...]plots_vert_selection=%7B%7D&plots_coloring_selection=%7B%7D (*GIPR*).

GLP1R-amplified breast-cancer samples (defined as GLP1R CNA = 2) showed 0 deaths among 27 patients with available survival records (0% death rate) versus 421 deaths among 2,964 (14.2%) non-GLP1R-amplified patient records (log-rank P = 0.11) (**Table S3**). In de-duplicated survival-linked breast-cancer cases, GLP1R copy-number states were centered on CNA = 0 (n = 665), with smaller groups at CNA = 1 (n = 236), CNA = -1 (n = 162), and CNA = 2 (n = 16), and crude death rates of 13.6%–14.2% across the CNA = -1, 0, and 1 groups versus 0% in the GLP1R-amplified subgroup (CNA=2).

In breast cancer cell-line screening experiments, there was no significant growth inhibition induced by the tested GLP-1 receptor agonists (liraglutide, BETP, geniposide) (**Figure S2**), CRISPR-mediated *GLP1R* knockout (**Figure S3a**), or RNA interference (RNAi)-mediated *GLP1R* knockdown (**Figure S3b**). Breast cancer cell lines generally show low GLP1R expression and near-neutral copy-number values, without evidence of a corresponding broad tumor-cell fitness dependency (**Figures S3c-d**).

*GIPR* alterations were observed in 61 of 4,726 (1.3%) profiled patients across the same 30 breast cancer studies. The majority of these alterations were also copy-number alterations, including amplifications of 46 of 3,724 (1.2%) profiled patients and deep deletions in 6 (0.16%) patients. Mutations in *GIPR* were similarly rare to *GLP1R* mutations, with mutations seen in only 9 of 2,311 (0.4%) profiled patients. Again, the distribution of mutations along the *GIPR* sequence were not clustered to suggest any recurrent alteration hotspots. In patients with *GIPR*-altered tumors, there was a trend toward inferior overall survival (p = 0.04, q = 0.08, **Figure 6b**). Similar to *GLP1R*, breast cancer cell lines showed generally near-neutral GIPR copy-number values with heterogeneous expression **(Figure S4**).

## Discussion

This real-world study suggests that in patients with pre-existing breast cancer, subsequent initiation of semaglutide was associated with reduced mortality compared to initiation of other anti-diabetic medications. Among these semaglutide-treated patients, increased survival was associated with a higher maximum dose of semaglutide during the first year of therapy but not with the magnitude of weight loss achieved during this same period. This raises the possibility of a functional dissociation between the substantial weight loss effect of GLP-1 receptor agonists and an association in breast cancer that is not fully explained by weight-loss magnitude alone. Interestingly, in the primary analyses presented here, the average time between breast cancer diagnosis and the first semaglutide prescription was approximately 6 years, implying that most of these patients were in remission at the time of semaglutide initiation. The results suggest a potential role for semaglutide in limiting the progression of existing disease, possibly including minimal residual disease (MRD). An ongoing phase 2 clinical trial of tirzepatide in obese patients with MRD+ early breast cancer will help to evaluate this question^14^, and similar studies utilizing semaglutide and/or other GLP-1 receptor agonists may be warranted.

One possible interpretation is that semaglutide exposure before oncology treatment may have altered host or tumor context in a way that favored subsequent treatment response, although this remains hypothetical. The secondary analysis in which survival was assessed after initiation of an oncology therapy corroborates the main findings and offers additional context. In these patients, semaglutide exposure occurred before initiation of the index oncologic therapy. Thus, the survival advantage seen under this analysis framework could reflect a priming effect of semaglutide, in which treatment with semaglutide triggers tumor or host alterations that facilitate a more favorable response to subsequent chemotherapy, targeted therapy, or immunotherapy.

The pattern seen here is directionally consistent with the evolving human literature on GLP-1 receptor agonists in oncology. In a large observational study of patients with type 2 diabetes, GLP-1 receptor agonists were associated with lower incidence of several obesity-associated cancers relative to insulin. However, this study did not find a significant reduction in the risk of postmenopausal breast cancer, there was no significant advantage over metformin for any of the cancers assessed^7^. More recently, another retrospective study in adults with obesity found a lower overall cancer incidence among GLP-1 receptor agonist users compared to non-users, in particular with reduced risks of endometrial cancer, ovarian cancer, and meningioma. There was a trend to lower risk of breast cancer, but this finding did not reach statistical significance (hazard ratio 0.86, 95% CI 0.71-1.03)^8^. A cohort study of older adults with cancer and type 2 diabetes further reported lower all-cause mortality among GLP-1 receptor agonist users than among DPP4 inhibitor users, including among breast cancer patients^9^. Of note, this study did not identify any difference in mortality among GLP-1 receptor agonist users compared to SGLT2 users^9^. Taken together, these studies suggest a potential role for GLP-1 receptor agonists in reducing cancer incidence and post-cancer diagnosis mortality, with mixed but overall favorable signals in breast cancer.

The lack of survival advantage seen in tirzepatide-treated patients compared with the pooled anti-diabetic comparator cohort should be interpreted with appropriate caution. Unlike semaglutide, tirzepatide engages not only the GLP-1 receptor but also the gastric inhibitory polypeptide receptor (GIPR)^15,16^. Thus, it is certainly feasible that these agents could have variable impacts on cancer outcomes despite their shared propensity for inducing weight loss and other favorable cardiometabolic rewiring^8,17–21^. However, the matched cohorts considered in this analysis were small, with 10 times fewer patients compared to the cohorts evaluated in the semaglutide versus pooled comparator analysis. Further, due to its later approval and corresponding shorter time on the market, the tirzepatide-treated cohort inherently has a shorter average follow-up duration. These factors reduce the power and sensitivity of the mortality analyses, so the lack of evidence for a survival benefit seen in breast cancer patients who subsequently initiated tirzepatide should not be interpreted as strong evidence for a lack of benefit. Indeed, the head-to-head matched analysis of tirzepatide and semaglutide in patients with pre-existing breast cancer did not reveal a significant survival difference, which could be compatible with non-superiority of one agent over the other in this setting.

The retrospective genomic and functional *in vitro* analyses do not demonstrate a convincing role for *GLP1R* or *GIPR* in the cell-intrinsic biology of breast cancer. Although *GIPR* genomic alterations were modestly associated with worse survival, the low alteration frequency and lack of recurrent “hotspot” mutations are less compatible with a role as a somatic driver of breast cancer. Increased expression of *GLP1R* and *GIPR* has been associated with improved survival in several tumor types including breast cancer^22^ (**Figure S5**), but these associations are not consistent across all studies and do not necessarily imply increased protein expression or receptor signaling that could contribute to altered tumor biology. Moreover, cell line perturbation screens showed that GLP1R or GIPR loss of function (LOF) did not significantly impact breast cancer cell viability, although it is possible that increased signaling through these receptors, as is induced by the therapies analyzed here, could impact tumor cell dynamics in ways that are not reflected in these LOF screens. In any case, there are other non-tumor intrinsic mechanisms by which GLP-1 receptor signaling could impact breast cancer biology, including rewiring of systemic host metabolism, altered tumor cell metabolism, or modulation of inflammatory pathways that contribute to anti-tumor immunity. In line with this, a recent study found that semaglutide reduced tumor growth and progression in a pre-clinical breast cancer model, possibly via immune reprogramming including increased dendritic cell maturation, decreased regulatory T cell abundance, and enhanced cytotoxic T cell function^23^.

This study has limitations. First, this was a retrospective study using real-world electronic health record (EHR) data and therefore is vulnerable to multiple sources of bias. Although propensity matching was performed to mitigate such bias to some extent, there are likely residual unmeasured confounding factors. For example, there may be selection bias in breast cancer patients who are selected to subsequently initiate semaglutide compared to metformin, SGLT2 inhibitors, or DPP4 inhibitors. Such a systematic difference could contribute to confounding in the survival analyses presented here. Second, the lack of consideration of cancer stage at the index dates (i.e. date of initiation of either semaglutide or an oncologic therapy) is a significant limitation, given that imbalance of stage would be expected to produce divergent survival curves irrespective of therapy. While cancer stage is not captured in easily accessible structured EHR data fields, curation of staging information from unstructured clinical notes would improve the robustness of the study. Third, the analyses that use the “landmark” framework incorporate a one-year period between semaglutide or tirzepatide initiation and the start of the follow-up period. This excludes patients who died or were lost to follow-up during the one-year pre-landmark period. While this same criteria was applied across all of the analyzed cohorts and thus does not introduce immortal time bias per se, there could be confounding factors introduced during those one year that impact the group assignment (i.e. maximum dose achieved or weight loss magnitude) and are not accounted for in the study design. Fourth, the study protocol did not match on exposure to specific oncologic therapies. The percentage of patients receiving different relevant classes of therapies (e.g., hormone therapy, HER2-directed therapy, CDK4/6 inhibitors, checkpoint blockade) was similar in the main analysis, but the secondary analysis that is anchored on the date of oncologic therapy initiation could benefit from more stringent class-based matching. Fifth, the clinical outcome that was analyzed was overall survival and metastasis. Consideration of more granular oncology-specific outcomes such as cancer-related death, progression-free survival, and disease-free survival would add value in future studies. Furthermore, the metastasis analyses presented here are limited, as they do not distinguish metastases by primary tumor of origin or timing relative to semaglutide initiation. Future studies should more rigorously adjudicate metastatic events using structured staging data and validated oncology endpoints. Finally, the AI-derived metastasis and LLM-derived cause-of-death analyses were exploratory and require explicit validation metrics and manual-review benchmarking.

Overall, these real-world analyses suggest a survival benefit for breast cancer patients who subsequently received semaglutide which was correlated with maximum attained dose but not weight loss magnitude. Taken together with other existing literature, these findings motivate prospective randomized studies to evaluate the impact of GLP-1 receptor agonists on diverse breast cancer-related outcomes across multiple therapeutic settings including as a neoadjuvant, combination, and/or adjuvant agent.

## Methods

### Study cohorts and exposure definitions

We assembled incident-user cohorts for semaglutide, tirzepatide, metformin, dipeptidyl peptidase 4 (DPP4) inhibitors, and sodium-glucose cotransporter 2 (SGLT2) inhibitors using harmonized medication records. The index date was defined as the date of the first qualifying prescription for the cohort-defining medication or medication class. Primary comparative analyses were anchored on semaglutide or tirzepatide as the exposure cohorts and compared with pooled or pairwise active-comparator cohorts constructed from metformin, DPP4 inhibitors, and SGLT2 inhibitors. Follow-up began on the index date and continued until death or the last observed clinical record.

### Breast-cancer ascertainment and baseline covariates

Patients were considered to have prior breast cancer only if they had at least 3 breast-cancer-coded diagnosis rows on at least 3 distinct dates before the diabetes-drug index date. We recorded the first qualifying preindex breast-cancer diagnosis date and used it to calculate time from qualifying prior breast cancer to index. Baseline demographic covariates included age at index and sex. Baseline body mass index (BMI) and baseline weight were defined as the nearest available measurements recorded from 365 days before through 14 days after index. Baseline type 2 diabetes was defined by at least 3 diagnosis-coded dates before index. We also derived the interval from the most recent mapped oncology treatment before index to the index date and categorized prior oncology-treatment history into endocrine therapy, chemotherapy, HER2-directed therapy, CDK4/6 inhibitor exposure, PARP inhibitor exposure, checkpoint immunotherapy, and other targeted therapy (**Table S1**). Pre-index breast cancer staging was derived via large language model (LLM) extraction from unstructured clinical notes in the 2-year window prior to the index date.

### Propensity-matched comparative survival analyses from drug index

For drug-index comparative analyses, propensity scores were estimated using logistic regression. Matching covariates were age at index, baseline BMI, baseline weight, index year, days from qualifying prior breast-cancer diagnosis to index, days from the most recent oncology treatment to index, sex, baseline type 2 diabetes, and the preindex oncology-treatment exposure flags described above. Patients were matched 1:1 without replacement by nearest-neighbor matching with a propensity-score caliper of 0.2 across all matching variables Near-exact matching was performed on index year. We performed pooled active-comparator analyses and separate pairwise analyses versus metformin, DPP4 inhibitors, and SGLT2 inhibitors. The primary outcome was all-cause mortality. Kaplan-Meier curves were displayed through 24 months, and patients were censored at the last observed clinical record.

### Analyses of metastatic disease burden and cause of death

To characterize post-treatment metastatic disease burden, two complementary approaches were employed. First, structured ICD diagnosis codes were queried across the propensity-matched cohorts to identify documented metastatic diagnoses in the post-index period. Second, an AI-based natural language processing (NLP) pipeline was applied to unstructured clinical notes using GPT-OSS-20B to extract and harmonize metastasis-related disease mentions, capturing diagnoses that may not be reflected in structured coding. For cause of death ascertainment, clinical notes from all deceased patients were processed exclusively using the GPT-OSS-20B model, which was prompted to identify and summarize the primary cause of death from available documentation. Extracted causes were subsequently harmonized into six high-level categories: cancer progression/metastasis, cardiac events, sepsis/septic shock, respiratory failure, hemorrhage/stroke, and other.

### Dose, weight-loss, and persistence one-year landmark analyses

Within semaglutide- and tirzepatide-treated cohorts, we performed landmark analyses at 1 year after drug initiation. Patients had to be alive and observable through the landmark to enter the corresponding post-landmark analysis. Maximum attained dose was defined as the highest recorded dose through the landmark. For semaglutide, exact dose bands were 0.25 mg, 0.5 mg, 1.0 mg, 1.7 mg, 2.0 mg, and 2.4 mg, and low versus high dose was defined as 0.25-1.0 mg versus 1.7 mg or greater. For tirzepatide, exact dose bands were 2.5 mg, 5.0 mg, 7.5 mg, 10.0 mg, 12.5 mg, and 15.0 mg, and low versus high dose was defined as 2.5-10.0 mg versus 12.5 mg or greater. Maximum weight loss was defined as the greatest percentage reduction from baseline weight to the lowest observed postindex weight before the landmark. Patients were categorized into prespecified weight-loss strata of less than 5%, 5-10%, 10-15%, 15-20%, and 20% or greater. Prescription persistence was evaluated from the interval between the last prescription date before the landmark and the landmark date; patients were classified as persistent if the last prescription occurred within 90 days of the landmark and otherwise as discontinued. Post-landmark mortality was then evaluated over the subsequent 24 months.

### Oncology-treatment-indexed analyses

To assess whether the survival association was preserved when follow-up was anchored to oncology treatment rather than to diabetes-drug initiation, we identified the first mapped oncology medication event occurring after the diabetes-drug index and reset the analysis index to that oncology-treatment date. We then repeated semaglutide active-comparator matching and survival analyses using the same covariates and censoring framework as in the primary drug-index analysis.

### Tumor-genomic and preclinical context analyses

To place the clinical findings in biological context, we performed exploratory breast-cancer tumor-genomic survival analyses for GLP1R and GIPR alteration status and examined GLP1R-related pharmacologic and dependency signals in DepMap breast-cancer cell lines. These analyses were conceptually distinct from the EHR-based treatment analyses and were intended as mechanistic context rather than as causal validation of the clinical associations.

### Statistical analysis

Continuous variables were summarized as means with standard deviations and categorical variables as counts and percentages. Survival comparisons were assessed with Kaplan-Meier estimation and log-rank testing. Comparative analyses reported matched cohort size, event counts, person-time, and 24-month Kaplan-Meier event probabilities. Statistical significance for metastasis outcomes and cause-of-death comparisons was assessed using chi-square p-values. No imputation was performed for missing data. The relationship between maximum attained dose and maximum percentage weight change was summarized with linear regression across dose groups. All analyses were observational and should be interpreted as hypothesis-generating rather than causal. Analyses were performed in Python 3.13.1 using pandas 3.0.0, NumPy 2.4.2, SciPy 1.17.0, and Matplotlib 3.10.8; propensity score modeling, survival analyses and single-cell expression profiling were implemented within the same analytic pipeline.

## Data Source

This study analyzed de-identified EHR data from academic medical centers in the United States via the nference nSights Analytics Platform. Prior to analysis, all data underwent expert determination de-identification satisfying HIPAA Privacy Rule requirements (45 CFR §164.514(b)(1)), employing a multi-layered transformation approach for both structured data (cryptographic hashing of identifiers, date-shifting, geographic truncation) and unstructured clinical text (ensemble deep learning and rule-based methods with >99% recall for personally identifiable information detection)^17,18^. nference established secure data environments within each participating center, housing these de-identified patient data governed by expert determination. These de-identified data environments were specifically designed to enable data access and analysis without requiring Institutional Review Board oversight, approval, or exemption confirmation. Accordingly, informed consent and IRB review were not required for this study.

## Data Availability

This study involves the analysis of de-identified Electronic Health Record (EHR) data via the nference nSights Federated Clinical Analytics Platform (nSights). Data shown and reported in this manuscript were extracted from this environment using an established protocol for data extraction, aimed at preserving patient privacy. The data has been de-identified pursuant to an expert determination in accordance with the HIPAA Privacy Rule. Any data beyond what is reported in the manuscript, including but not limited to the raw EHR data, cannot be shared or released due to the parameters of the expert determination to maintain the data de-identification. The corresponding author should be contacted for additional details regarding nSights.

## De-identification and HIPAA compliance certification

Prior to analysis, all EHR data were de-identified under an expert determination consistent with the Health Insurance Portability and Accountability Act (HIPAA) Privacy Rule (45 CFR §164.514(b)(1)). The de-identification methodology employed a multi-layered transformation approach to both structured and unstructured data fields^24^. In structured data, direct identifiers including patient names and precise geographic locations were excluded entirely, while indirect identifiers underwent specific transformations: patient identifiers, medical record numbers, and accession numbers were replaced with one-way cryptographic hashes using confidential salts to preserve linkage across patient encounters; all dates were shifted backward by patient-specific random offsets (1–31 days) to preserve temporal relationships while obscuring exact event timing; the ZIP codes were truncated to two-digit state-level resolution; and continuous variables including age, height, weight, and body mass index were thresholded to prevent identification of extreme values (for example, ages ≥89 years transformed to ‘89+’ and BMI >40 transformed to ‘40+’). In unstructured clinical text, an ensemble de-identification system that combines attention-based deep learning models with rule-based methods achieved an estimated >99% recall for personally identifiable information (PII) detection, with detected identifiers replaced by plausible fictional surrogates^24^.

## Data Harmonization

To address heterogeneity in EHR data, we harmonized clinical variables including medications, anthropometric measurements, and diagnoses to standardized concepts. For medications, we first constructed a standardized drug concept database combining the nSights knowledge graph with RXNorm (https://www.nlm.nih.gov/research/umls/rxnorm/index.html) hierarchies to capture ingredient, brand, and dose-specific information^25^. EHR medication records were matched using a hierarchical approach prioritizing RXNorm codes when available, followed by ingredient-level matching, and finally natural language processing and pattern matching on free-text medication orders when structured codes were absent. For anthropometric measurements (height, weight, BMI), we created a unified vocabulary from SNOMED (https://www.snomed.org/, https://athena.ohdsi.org) and LOINC (https://loinc.org/) terminologies and matched EHR measurement descriptions using standardized text matching algorithms with abbreviation expansion and synonym resolution; ambiguous mappings were resolved using OpenAI GPT-4o (https://platform.openai.com/docs/models/gpt-4o) with summary statistics as context, followed by manual verification. For diagnoses, we developed a hierarchical disease concept database from the nSights knowledge graph and matched EHR diagnosis descriptions and codes by identifying the most specific common child concept in the hierarchy. This approach enabled consistent identification of clinical entities while preserving granularity where available.

## Code Availability

The analysis code is not publicly available. The corresponding author should be contacted for additional details.

## Author Contributions

K.M. and V.S. conceived and designed the study. K.M. conducted data queries, large language model (LLM) extraction, and statistical analyses. A.J. conducted molecular data analyses. All authors contributed to the interpretation of results and the writing of the manuscript.

## Acknowledgements

We thank the nference engineering team for the development of the nSights federated AI platform. We also thank Patrick Lenehan for the critical study review and Gourab Saha for genomics data input.

## SUPPLEMENTARY MATERIAL

**Table S1.**
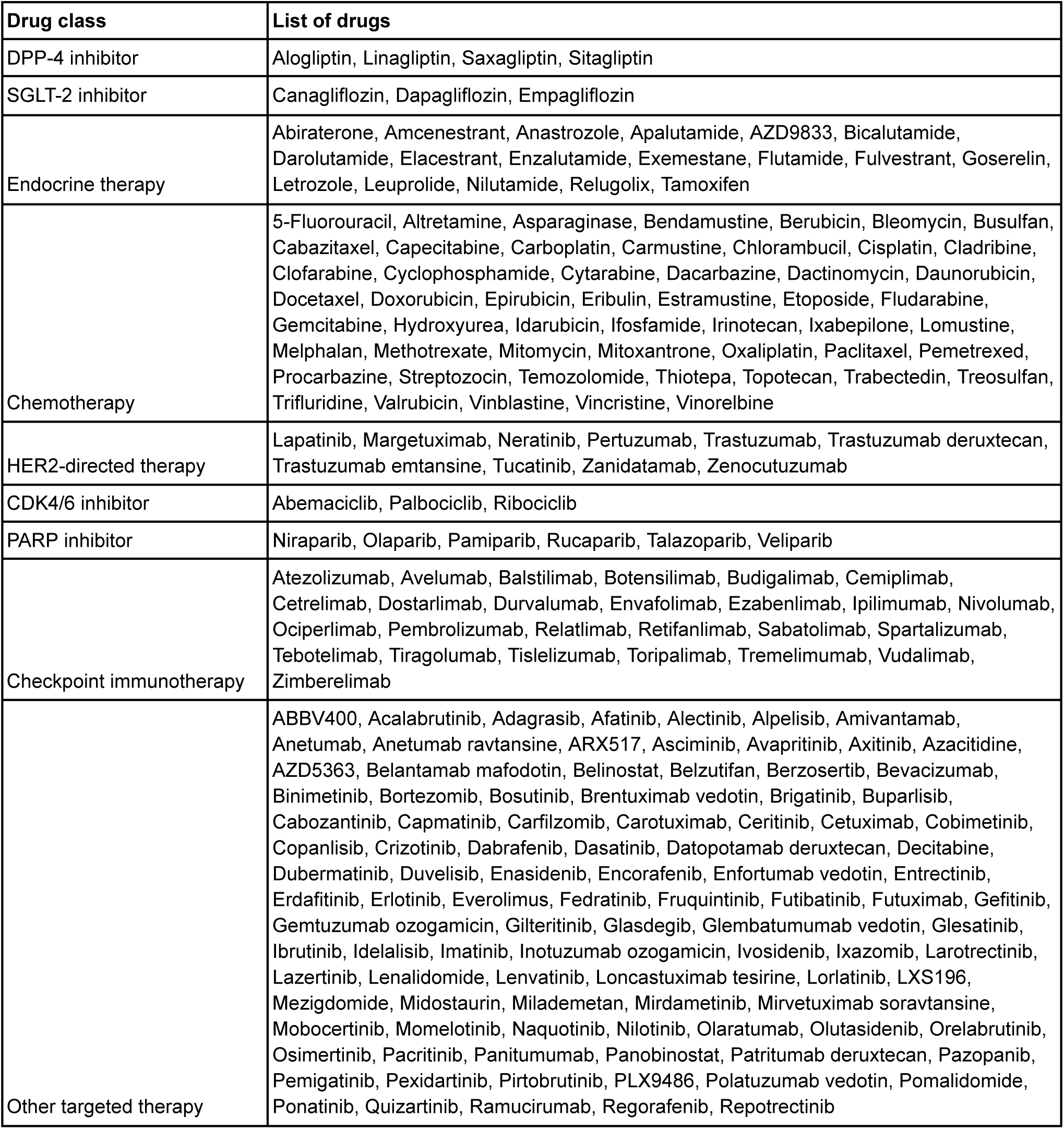
Drug-class definitions used in the breast-cancer analyses, listing the constituent DPP-4 inhibitor and SGLT-2 inhibitor comparator drugs and the canonical oncology agents grouped into the treatment-history classes used for breast-cancer covariate adjustment.

**Table S2.**
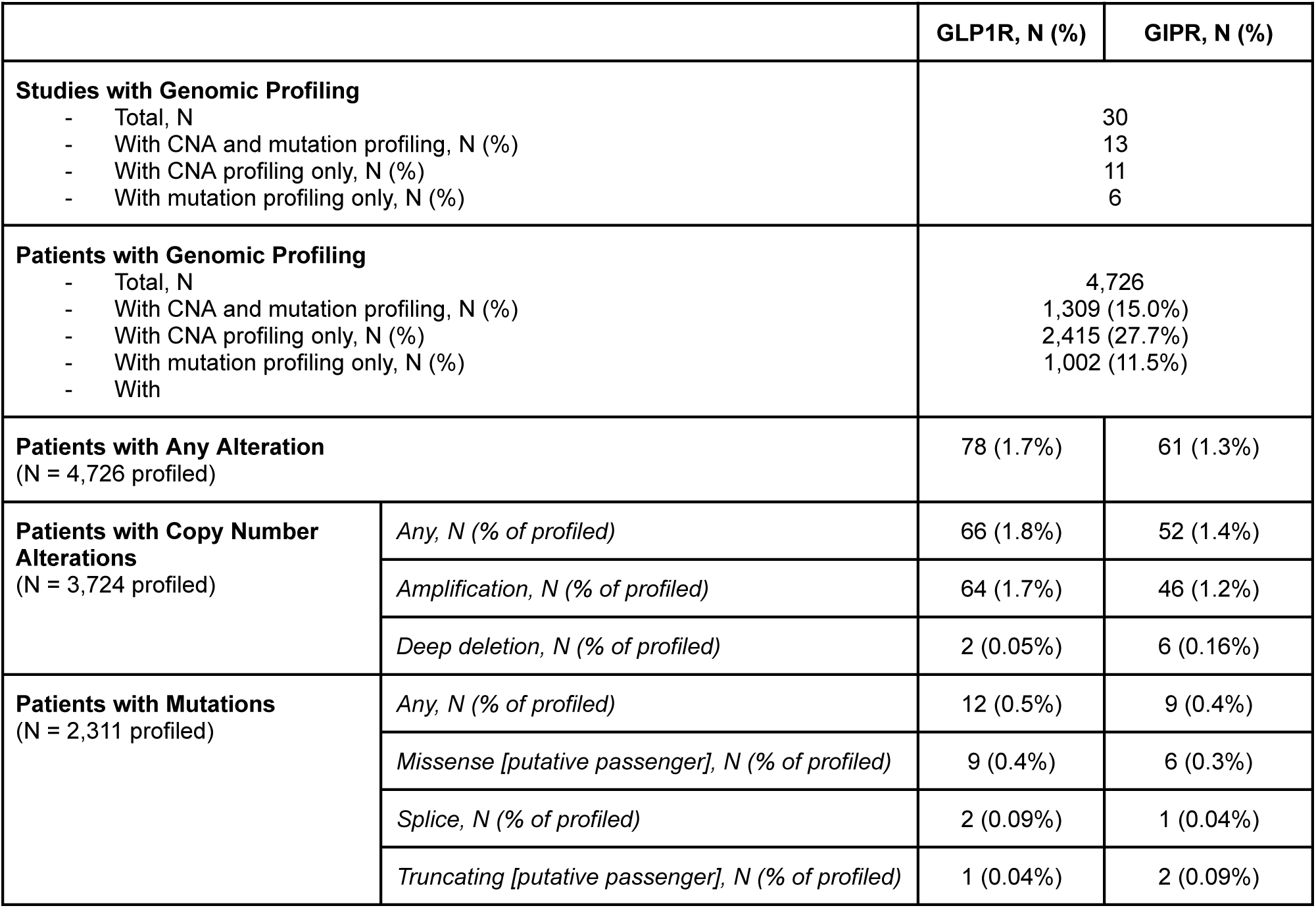
Summary of GLP1R and GIPR genomic alterations across breast cancer studies. Data was accessed from the cBioPortal Portal For Cancer Genomics, with tabular data downloaded from the OncoPrint tab. The specific alterations types (amplification, deep deletion, missense, splice, and truncating) are provided by the cBioPortal output file. The label of “putative passenger” mutations is based on prior curated knowledge from OncoKB and statistically unexpected recurrence as defined by the Cancer Hotspots database. The URL for data download can be accessed at: www.cbioportal.org/results/oncoprint?comparison_selectedGroups=%5B%22GLP1R%20unaltered%22%2C%22Unaltered%20group%22%2C%22GIPR%22%5D&mutations_gene=GIPR&tab_index=tab_visualize&Action=Submit&session_id=69e902c1d0c16d54f08676c1&plots_horz_selection=%7B%7D&plots_vert_selection=%7B%7D&plots_coloring_selection=%7B%7D.

**Table S3.** Copy number alteration (CNA) for GLP1R in breast cancer samples.

| GLP1R CNA category | Patients | Deaths | Death rate |
| --- | --- | --- | --- |
| -1 | 162 | 23 | 14.20% |
| 0 | 665 | 93 | 14.00% |
| 1 | 236 | 32 | 13.60% |
| 2 | 16 | 0 | 0.00% |

**Figure S1.**
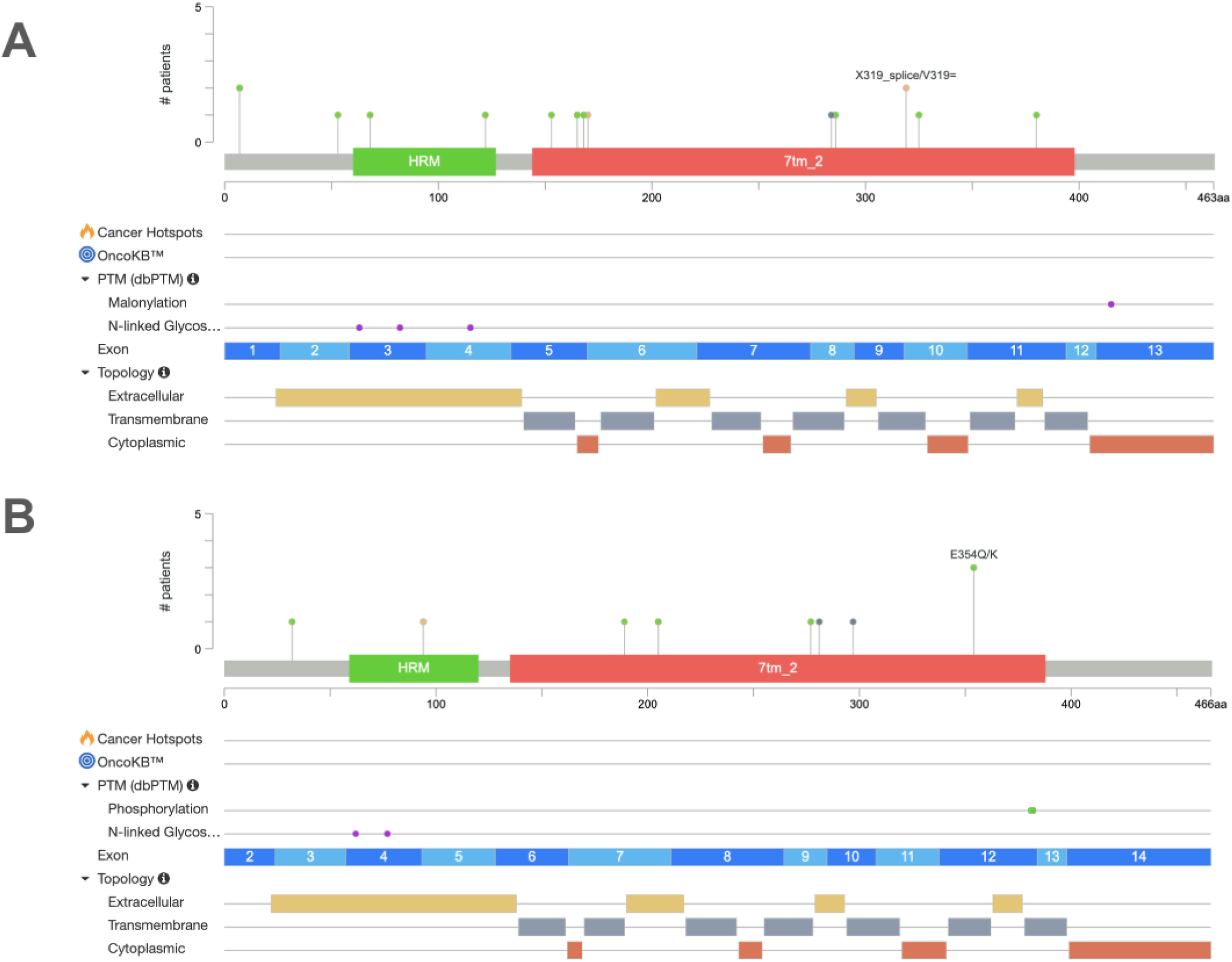
Lollipop visualizations of the *GLP1R (A)* and *GIPR (B)* mutation landscapes in breast cancer. Data are sourced from cBioPortal for Cancer Genomics (https://www.cbioportal.org/results/mutations?comparison_selectedGroups=%5B%22GLP1R%20unaltered%22%2C%22GLP1R%22%2C%22Unaltered%20group%22%5D&mutations_gene=GLP1R&tab_index=tab_visualize&Action=Submit&session_id=69e902c1d0c16d54f08676c1&plots_horz_selection=%7B%7D&plots_vert_selection=%7B%7D&plots_coloring_selection=%7B%7D and https://www.cbioportal.org/results/mutations?comparison_selectedGroups=%5B%22GLP1R%20unaltered%22%2C%22GLP1R%22%2C%22Unaltered%20group%22%5D&mutations_gene=GIPR&tab_index=tab_visualize&Action=Submit&session_id=69e902c1d0c16d54f08676c1&plots_horz_selection=%7B%7D&plots_vert_selection=%7B%7D&plots_coloring_selection=%7B%7D).

**Figure S2.**
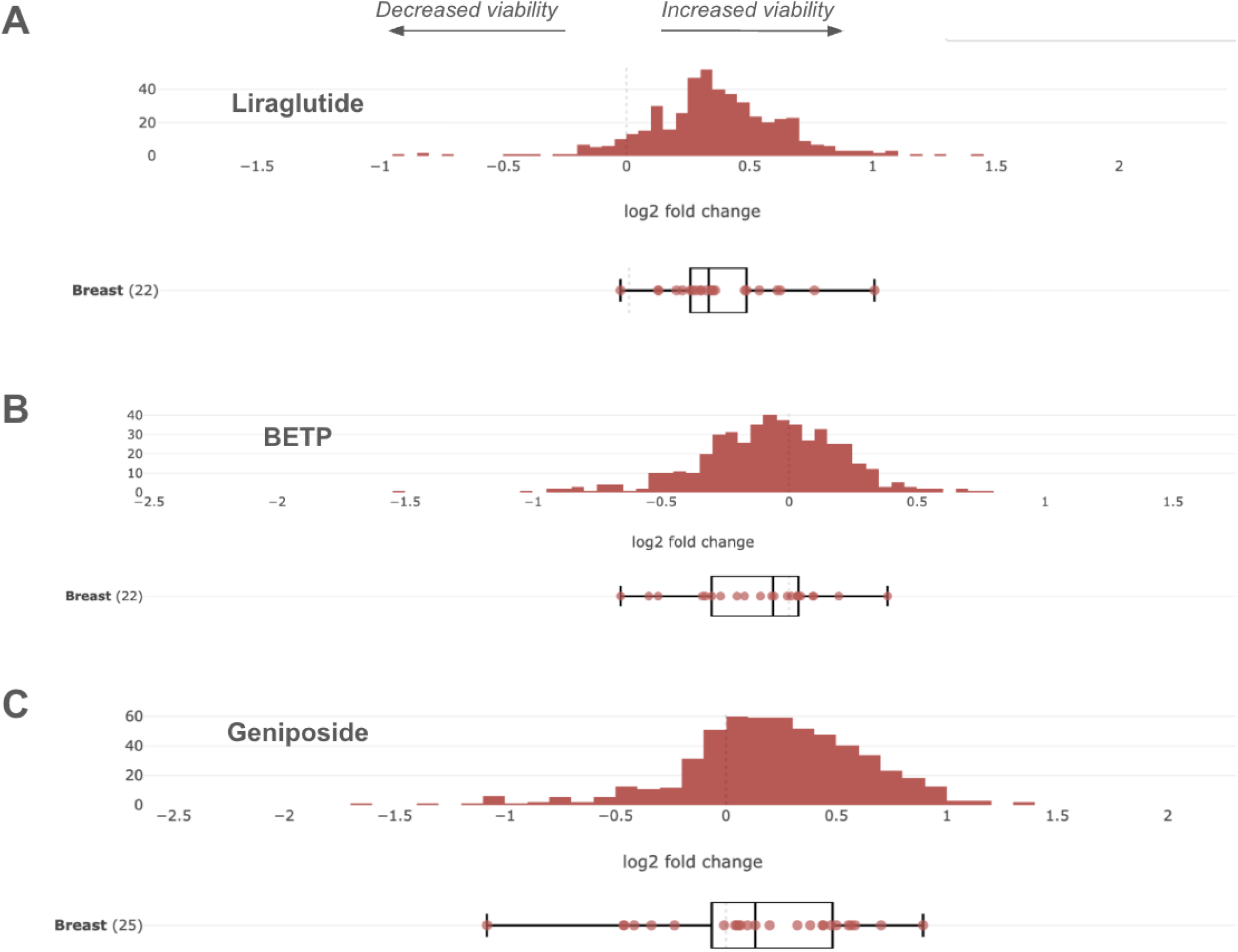
Impact of GLP-1 receptor agonists on cancer cell line viability in the PRISM Repurposing Primary dataset. Histogram of log2 fold-change values represents the distribution of PRISM viability responses to **(A)** Liraglutide, **(B)** BETP, and **(C)** geniposide across cell lines of all assayed cancer lineages in the PRISM dataset. The distribution of viability responses for 22 breast cancer cells lines to each compound is shown as a box plot with overlaid points representing each individual cell line. Data is accessed from the Broad Institute DepMap portal (https://depmap.org/portal/compound/LIRAGLUTIDE?tab=dependency, https://depmap.org/portal/compound/BETP?tab=dependency, https://depmap.org/portal/compound/GENIPOSIDE?tab=dependency).

**Figure S3.**
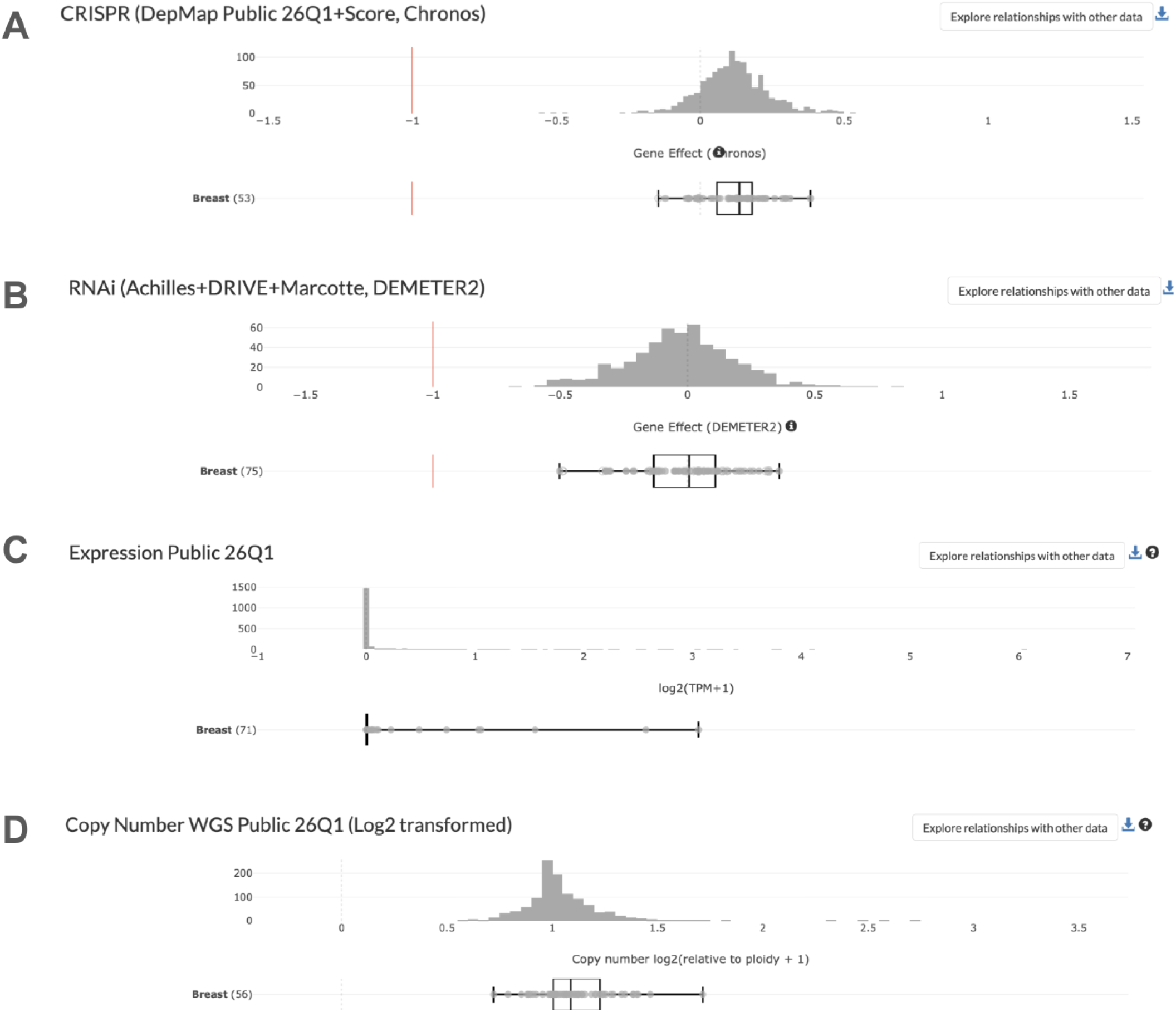
Summary of GLP1R dependency, expression, and copy number status across cell lines from the Broad Institute DepMap. **(A)** Impact of CRISPR-mediated GLP1R knockout on cell line viability. (B) Impact of RNAi-mediated GLP1R knockdown on cell line viability. (C) Distribution of GLP1R mRNA expression, expressed as log2(TPM+1). (D) Distribution of GLP1R copy number status. In each panel, the histogram depicts the distribution of the corresponding metric across all cell lines of all assayed cancer lineages, and the boxplot with overlaid points depicts the distribution of the corresponding metric across all assayed breast cancer cell lines. Data was accessed from: https://depmap.org/portal/gene/GLP1R?tab=dependency.

**Figure S4.**
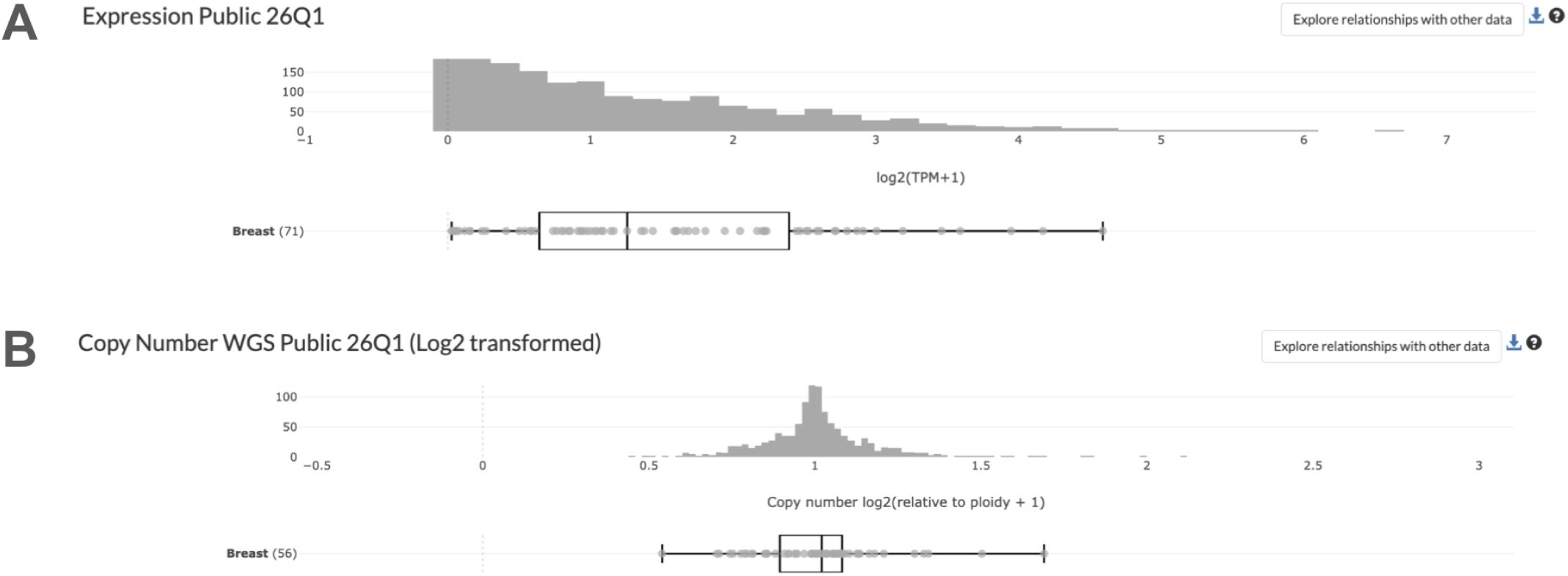
Summary of *GIPR* expression and copy number status across cell lines from the Broad Institute DepMap. (A) Distribution of GLP1R mRNA expression, expressed as log2(TPM+1). (B) Distribution of GLP1R copy number status. In each panel, the histogram depicts the distribution of the corresponding metric across all cell lines of all assayed cancer lineages, and the boxplot with overlaid points depicts the distribution of the corresponding metric across all assayed breast cancer cell lines. Data was accessed from: https://depmap.org/portal/gene/GIPR?tab=characterization&dependency=RNAi_Ach&characterization=expression and https://depmap.org/portal/gene/GIPR?tab=characterization&dependency=RNAi_Ach&characterization=copy_number_relative.

**Figure S5.**
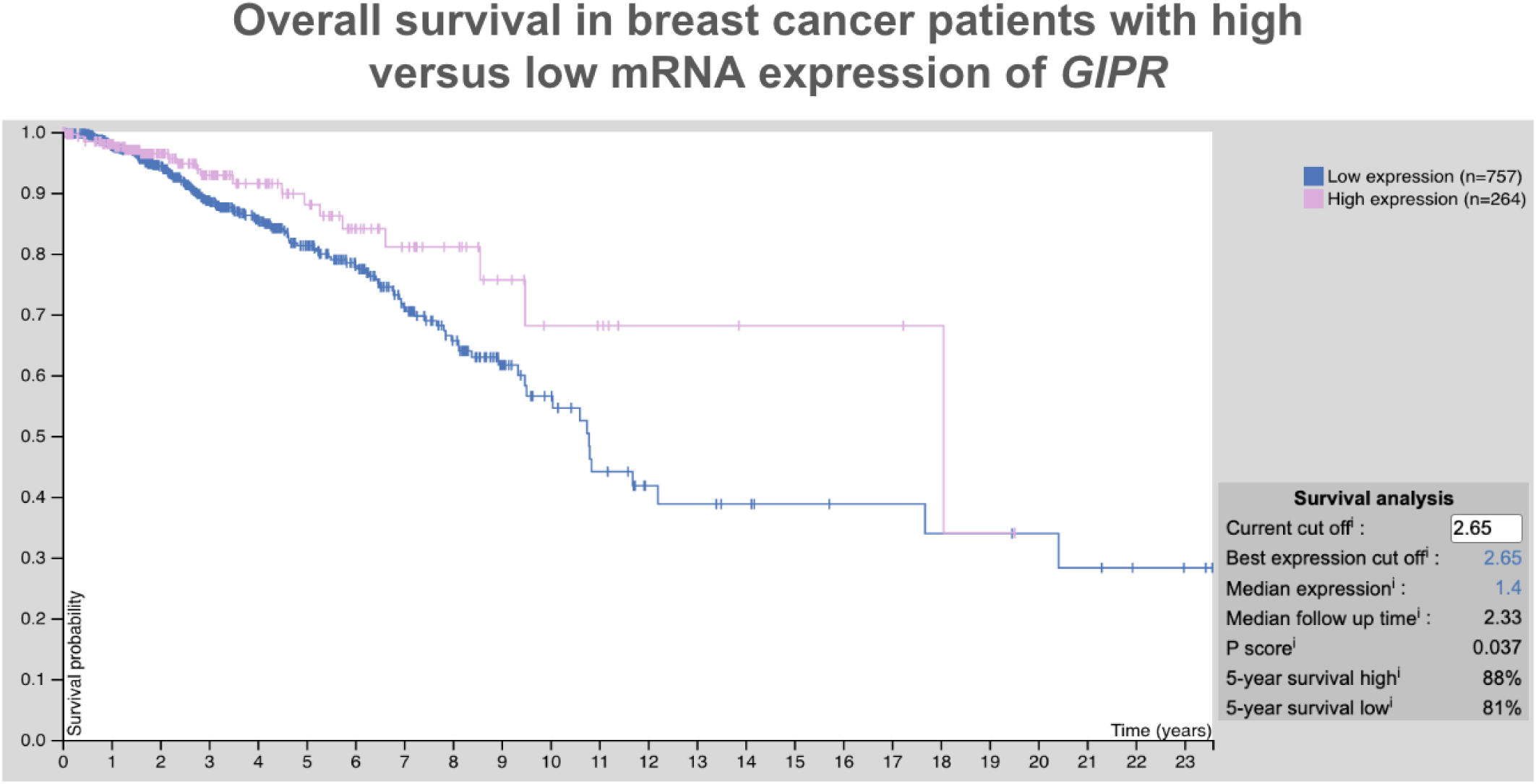
Overall survival of breast cancer patients stratified by *GIPR* mRNA expression levels. Data is derived from 1,021 breast invasive carcinoma patients in The Cancer Genome Atlas (TCGA). Visualization and statistics are provided on the Human Protein Atlas portal, with an optimized expression threshold to maximize separation of survival curves between the high- and low-expressing groups (https://www.proteinatlas.org/ENSG00000010310-GIPR/cancer/breast+cancer#BRCA_validation).

